# Bidirectional Association and Genetic Background of Anxiety Disorders and General Medical Conditions

**DOI:** 10.1101/2025.10.10.25337730

**Authors:** Anniina Tervi, Eos Suomalainen, Heidi Perälampi, Nelli Frilander, Joonas Naamanka, FinnGen, Anxiety Disorders Working Group of the Psychiatric Genomics Consortium, Jaana Suvisaari, Marko Elovainio, Iiris Hovatta

## Abstract

**Importance:** Anxiety disorders are often comorbid with general medical conditions, but the extent and directionality of these associations and shared genetic risk factors are not well understood.

**Objective:** To investigate the potential associations and the directionality between anxiety disorders and general medical conditions, and to explore the underlying shared genetic risk.

**Design:** We used FinnGen data since the beginning of the registries or in Cox regression analyses between 2011 and 2022.

**Setting:** The longitudinal FinnGen study combines electronic health record and genomic data.

**Participants:** Over 500,000 FinnGen participants.

**Exposures:** Anxiety disorders and general medical conditions were defined based on the International Classification of Diseases codes. Anxiety disorders included panic disorder, generalized anxiety disorder, and phobias.

**Main Outcomes and Measures:** We conducted Cox proportional hazard analyses to examine associations between anxiety disorders and 11 general medical condition broad categories. Linkage disequilibrium score regression was used to assess shared genetic risk and two-sample Mendelian randomization to estimate causality using publicly available genome-wide association study summary statistics alongside FinnGen data.

**Results:** We observed bidirectional associations between anxiety disorders and most general medical conditions. Gastrointestinal and neurological diseases showed a particularly strong bidirectional association. Prior gastrointestinal (HR [CI 95%] = 2.02 [1.97-2.08]) and neurological (HR [CI 95%] = 1.96 [1.91-2.01]) diseases strongly predicted subsequent anxiety disorders. Conversely, a prior anxiety disorder diagnosis predicted later gastrointestinal (HR [CI 95%] = 1.77 [1.73-1.82]) and neurological (HR [CI 95%] = 1.72 [1.68-1.76]) diagnosis. We found shared genetic risk (P < 2x10^-5^), notably between anxiety disorders and several gastrointestinal diseases (r_g_ = 0.41-0.71). Pleiotropy analysis highlighted regulatory regions in the genome as the main shared feature between anxiety disorders and gastrointestinal diseases as well as genes linked to brain function. Mendelian randomization suggested a causal association from anxiety disorders to asthma and gastrointestinal diseases, but not vice versa.

**Conclusions and Relevance:** The bidirectional association and shared genetic risk between anxiety disorders and general medical conditions suggest shared underlying pathophysiological mechanisms. The observed links, particularly the causal association from anxiety to certain medical conditions, emphasize the importance of considering somatic health in psychiatric care and vice versa.

## Introduction

The most recent Global Burden of Disease Study^1^ listed anxiety disorders as one of the leading causes with the highest incidence and years lived with disability. Their age-standardized disability-adjusted life-year rates from 2010 to 2021 were among the most significantly increased.^1^ Anxiety disorders often co-occur with general medical conditions (GMCs)^2–4^ but many features of these associations, including the underlying mechanisms, directionality, and long-term trajectories, remain largely unknown.

Genetic epidemiological studies over the past decade have found shared biological mechanisms underlying different medical conditions.^5^ These studies have showed the underlying genetic component and moderate heritability of anxiety disorders (single nucleotide polymorphism (SNP) based heritability 10–28%^6^) highlighting the underlying neurobiological intricacies in anxiety disorders.^7^ The most recent large-scale genome-wide association study (GWAS) with over 122,000 cases identified 58 independent genetic variants associated with anxiety disorders.^6^ The studies investigating shared genetic risks between anxiety disorders and GMCs have shown at least a moderate genetic correlation, for instance, with insomnia^8,9^, endometriosis,^9,10^ and pain^9^. Despite the progress in the field, the application of systematic and finely resolved definitions of anxiety to explore the underlying genetic overlap with GMCs is limited.

In this study, we used longitudinal electronic health record (EHR) and genetic data from the FinnGen study to explore the associations and underlying biological mechanisms between anxiety disorders and GMCs in over 500,000 individuals. Our particular interest was to investigate whether higher risk for anxiety disorders can be witnessed with a prior GMC or whether prior anxiety disorder increases the risk of subsequent GMC. In addition, we examined the shared genetic risk factors between anxiety disorders and GMCs alongside estimating the causality with genetic instruments.

## Methods

### Sample and Phenotypes

The FinnGen initiative is a large-scale public-private partnership in Finland that combines genomic data with extensive EHR data from approximately 500,000 participants.^11^ Our study used FinnGen data release 12 to define phenotypes for both epidemiological and genetic analyses (eTables 1-4). We used ICD (8th, 9th, and 10th revisions) and KELA (Social Insurance Institution of Finland) codes from primary care, inpatient, outpatient, death, and drug purchase registries.

Anxiety disorders were treated as a single category, including phobic anxiety (F40) and other anxiety disorders (F41) from ICD-10 (eTables 1,3). We did not include disorders mainly affecting childhood (separation anxiety disorder and selective mutism), or those caused by substances or other medical conditions. Obsessive-compulsive disorder (OCD) and post-traumatic stress disorder (PTSD) were also not included to focus specifically on core anxiety disorders.

GMCs were grouped into 11 broad categories based on ICD codes and prior research^2,3,12^ (eTable 1). We excluded individuals with certain conditions in epidemiological analyses such as autism and schizophrenia due to FinnGen’s enriched specialized sample cohorts^11^ (eTable 2). We also excluded somatoform disorders to focus on genuine co-occurring GMCs (eTable 2).

For genetic analyses, we included all individuals without exclusions in GMCs (eTable 1) and created five custom anxiety definitions (eTable 3) to define cases and controls. We also utilized summary statistics without FinnGen data from the Psychiatric Genomics Consortium (PGC) Anxiety Disorders Working Group^6^ and publicly available GMC data (eTables 5,6).

### Epidemiological Analyses

We examined the prevalence of 11 broad GMC categories among individuals with anxiety disorders (Figure 1, eTable 7). This was compared to a control group, matched for age (+/- 2 years) and sex, that with 1:2 ratio. The Chi-squared test was used to determine statistical significance (P < 0.05).

**Figure 1.**
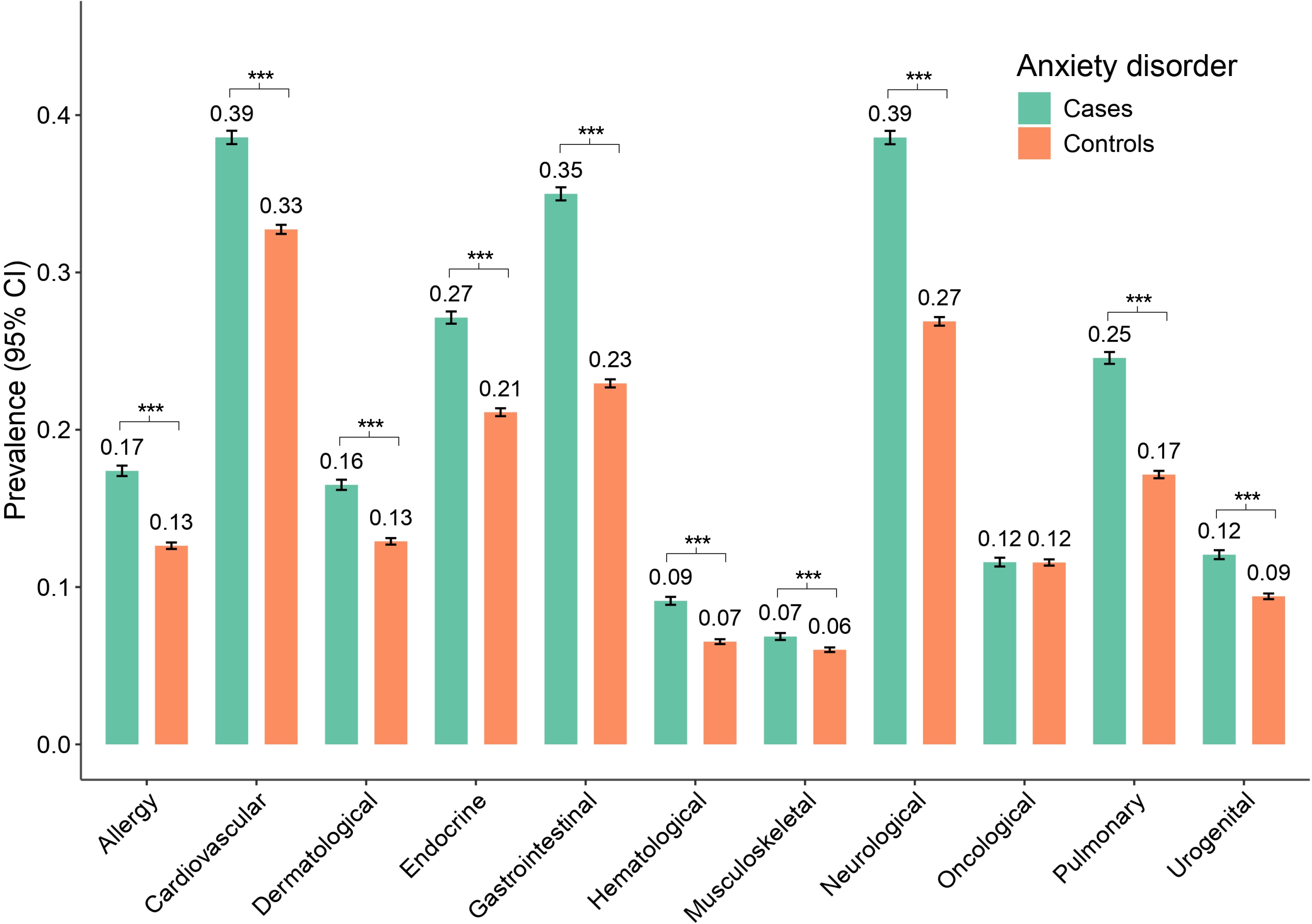
Prevalence of broad categorical general medical condition diagnoses in anxiety disorder cases and controls. Prevalence is shown with 95% confidence intervals (CIs) on the y-axis and broad categorical general medical conditions are listed on the x-axis. Control group was matched by age (+/- 2 years) and sex and consisted of twice the number of individuals as the anxiety disorder group. Significance of the differences in proportions was tested using χ2-test. Statistically significant (P < 0.001***) differences were observed in all broad categories except in oncological diseases. Created with BioRender.com.

We used multivariate Cox Proportional Hazards models^13^ to assess the bidirectional associations between anxiety disorders and GMCs, with age as the underlying time scale. The follow-up period was 12 years. All models were adjusted for sex, date of entry, and preceding comorbidities (GMCs and psychiatric conditions) (eTable 8). We created four models: unadjusted, adjusted for both GMCs and psychiatric conditions, adjusted for GMCs only, and adjusted for psychiatric conditions only.

We furthermore conducted time-dependent Cox Proportional Hazards models to evaluate how hazard ratios (HRs) changed with varying exposure durations to a preceding condition. The exposure was divided into seven intervals, from 0–6 months to ≥ 15 years.

### Genetic Analyses

FinnGen genotyping was performed using Affymetrix and Illumina arrays, with data imputed using the SiSu v4 reference panel.^11,14^ Quality control steps excluded individuals and variants with low genotyping quality.^11^ GWAS were conducted using the REGENIE pipeline^15^, adjusted for age, sex, genotyping batches, and genetic principal components.

We used linkage disequilibrium score regression (LDSC)^16^ to analyze the genetic correlation between anxiety disorders and GMCs. Initially, we used custom FinnGen GWAS summary statistics, and for sensitivity analyses, we additionally used publicly available data^17–30^ (eTables 5,6). Sensitivity analyses were performed for GMCs with a genetic correlation (r_g_) greater than 0.3 in the initial analyses.

To estimate causality, we used Mendelian Randomization (MR) with lead SNPs from anxiety disorder and GMC GWAS using R package, TwoSampleMR^31,32^. Egger intercept method was used to check for horizontal pleiotropic effects.

We further investigated the pleiotropic effects in genetic correlation using the CPASSOC^33^ method, which compares Z-scores from GWAS accounting for sample overlap. We ran both the SHom (homogeneous) and SHet (heterogeneous) models. We specifically focused on anxiety disorders and gastrointestinal diseases (irritable bowel syndrome, IBS, and gastro-esophageal reflux disease, GERD) that had a consistent r_g_ > 0.3 in LDSC. We also performed meta-analyses for these gastrointestinal diseases using METAL^34^ to increase power.

Detailed methods are reported in eMethods in Supplement 1.

## Results

### Prevalence

There were clear differences (P < 0.001) in prevalences of all broad categories, except oncological diseases, between individuals diagnosed with anxiety disorders and the control group, with anxiety disorder cases having higher prevalence (Figure 1, eTable 7). The most pronounced differences in prevalence between anxiety disorder cases and controls were observed in the neurological (χ² = (1, N = 151209) = 2096.6, P < 2.2x10^-16^) and gastrointestinal (χ² = (1, *N* = 151209) = 2494.7, P < 2.2x10^16^) disease categories (Figure 1, eTable 7).

### Bidirectional Relationship

We observed the strongest associations, using Cox regression analyses on the broad categorical level (adjusting for the date of entering to follow-up, sex, GMC and psychiatric comorbidities), with prior anxiety disorders and later gastrointestinal (HR [CI 95%] = 1.77 [1.73-1.82]), hematological (HR [CI 95%] = 1.84 [1.75-1.92]) and neurological diseases (HR [CI 95%] = 1.72 [1.68-1.76]) (Figure 2A, eTable 9, eFigures 1-6). In the other direction, prior gastrointestinal (HR [CI 95%] = 2.02 [1.97-2.08]) and neurological (HR [CI 95%] = 1.96 [1.91-2.01]) diseases and later development of anxiety disorders (Figure 2A, eTable 9, eFigure 1-6) had the strongest association. All HRs specified here were statistically significant with a P < 2x10^-16^.

**Figure 2.**
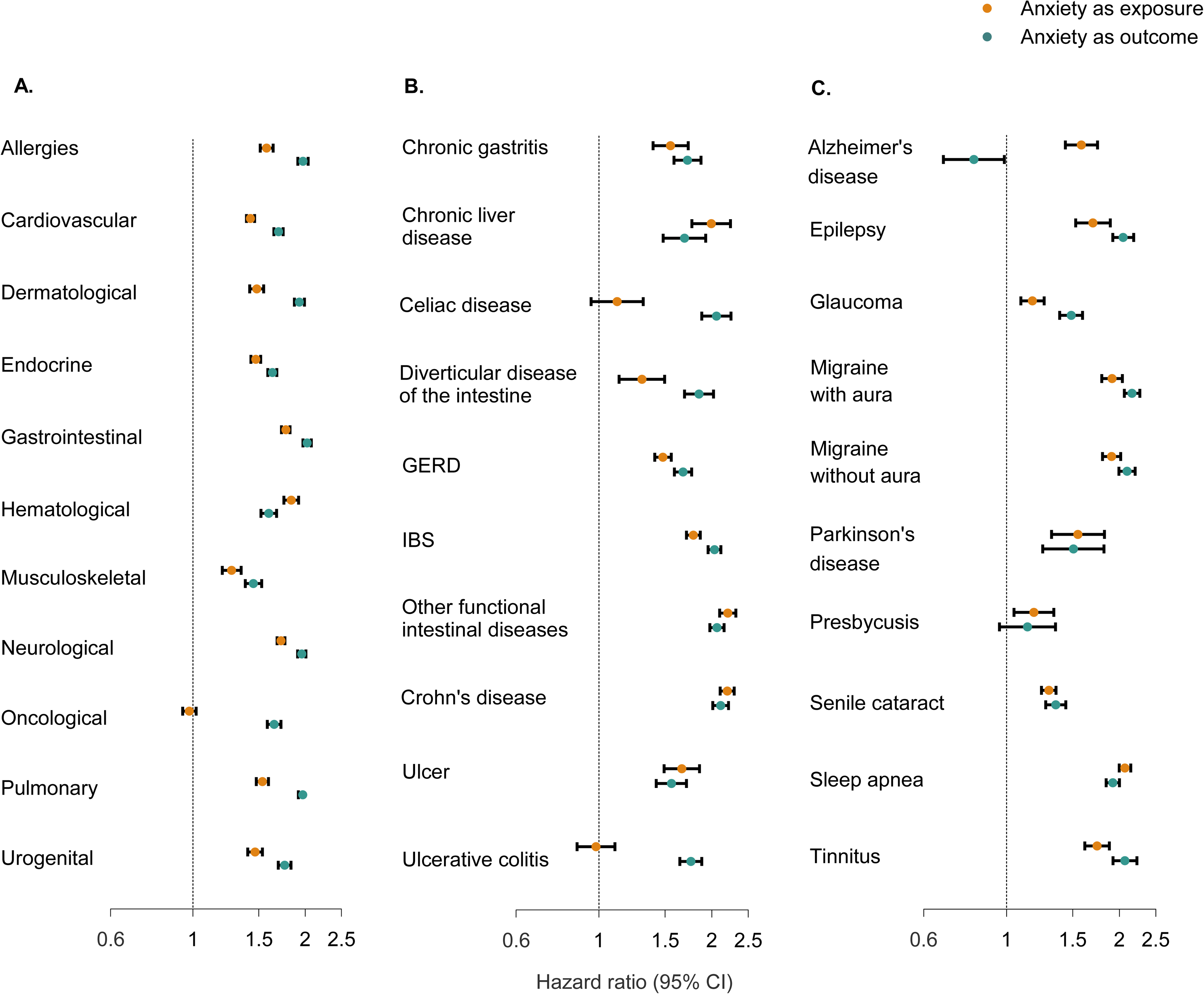
Bidirectional associations between anxiety disorders and general medical conditions. Cox proportional hazards models between anxiety disorders and general medical conditions (GMCs). Orange color depicting anxiety disorders as the exposure and teal color anxiety disorders as the outcome. GMCs are listed on the y-axis and hazard ratios with 95% confidence intervals (CIs) on the x-axis. Anxiety disorders bidirectional relationship with **A.** GMCs combined into broad categories, **B.** individual gastrointestinal diseases, and **C.** individual neurological diseases. The results shown here are adjusted for sex, the date of entry into follow-up, and other GMC and broad psychiatric disorder categories (model B, see eTables 9-10 for the results of the other models). IBS: Irritable bowel syndrome, GERD: gastro-esophageal reflux.

Overall, all gastrointestinal diseases were significantly associated (P < 0.001) with later anxiety disorders (Figure 2B, eTable 10, eFigure 2). We observed the strongest associations to both directions with IBS (anxiety as an exposure: HR [CI 95%] = 2.21 [2.10-2.32]; anxiety as the outcome: HR [CI95%] = 2.06 [1.98-2.16]) and other functional intestinal disorders (anxiety as an exposure: HR [CI 95%] = 2.20 [2.11-2.30]; anxiety as the outcome: HR [CI 95%] = 2.11 [2.01-2.22]) (Figure 2B, eTable 10, eFigure 2) all with a P < 2x10^-16^.

All neurological diseases, except Alzheimer’s disease (HR [CI 95%] = 0.82 [0.68-0.99], P = 0.04) and presbycusis (HR [CI 95%] = 1.14 [0.96-1.35], P= 0.15) were associated with later anxiety disorder, and anxiety disorder diagnosis was associated with all subsequent neurological diseases studied (Figure 2C, eTable 10, eFigure 3). For anxiety as the outcome, we found the strongest associations with migraine with aura (HR [CI 95%] = 2.16 [2.06-2.27]) and migraine without aura (HR [CI 95%] = 2.10 [1.99-2.20]) (Figure 2C, eTable 10, eFigure 3) both with a P < 2x10^-16^. With anxiety disorders as the exposure, we observed the strongest associations with migraine without aura (HR [CI 95%] = 1.91 [1.80-2.02]) and sleep apnea (HR [CI 95%] = 2.07 [2.00-2.14]) (Figure 2C, eTable 10, eFigure 3) all with a P < 2x10^-16^.

### Bidirectional Temporal Patterns

When examining time-dependent associations in the broad ICD-categories, we observed the strongest associations mostly within the first 6 months of exposure to the preceding condition (Figure 3, eTables 11-12, eFigures 7-9). In the models adjusted with comorbid conditions (model B) (Figure 3A, eTable 11), however, there was an increasing trend over time in all HRs where the strongest associations were mostly observed after ≥ 15 years of exposure (Figure 3A). The strongest association was between prior gastrointestinal (HR [CI 95%] = 2.87 [2.73-3.02]), dermatological (HR [CI 95%] = 2.85 [2.70-3.00]), allergic (HR [CI 95%] = 2.82 [2.64-3.01]) and neurological (HR [CI 95%] = 2.79 [2.64-2.95]) diseases after ≥ 15 years of exposure (Figure 3A, eTable 11). With anxiety as the exposure, the strongest association was in the category of hematological (HR [CI 95%] = 2.91 [2.64-3.21]) diseases after ≥ 15 years of exposure (Figure 3A, eTable 11).

**Figure 3.**
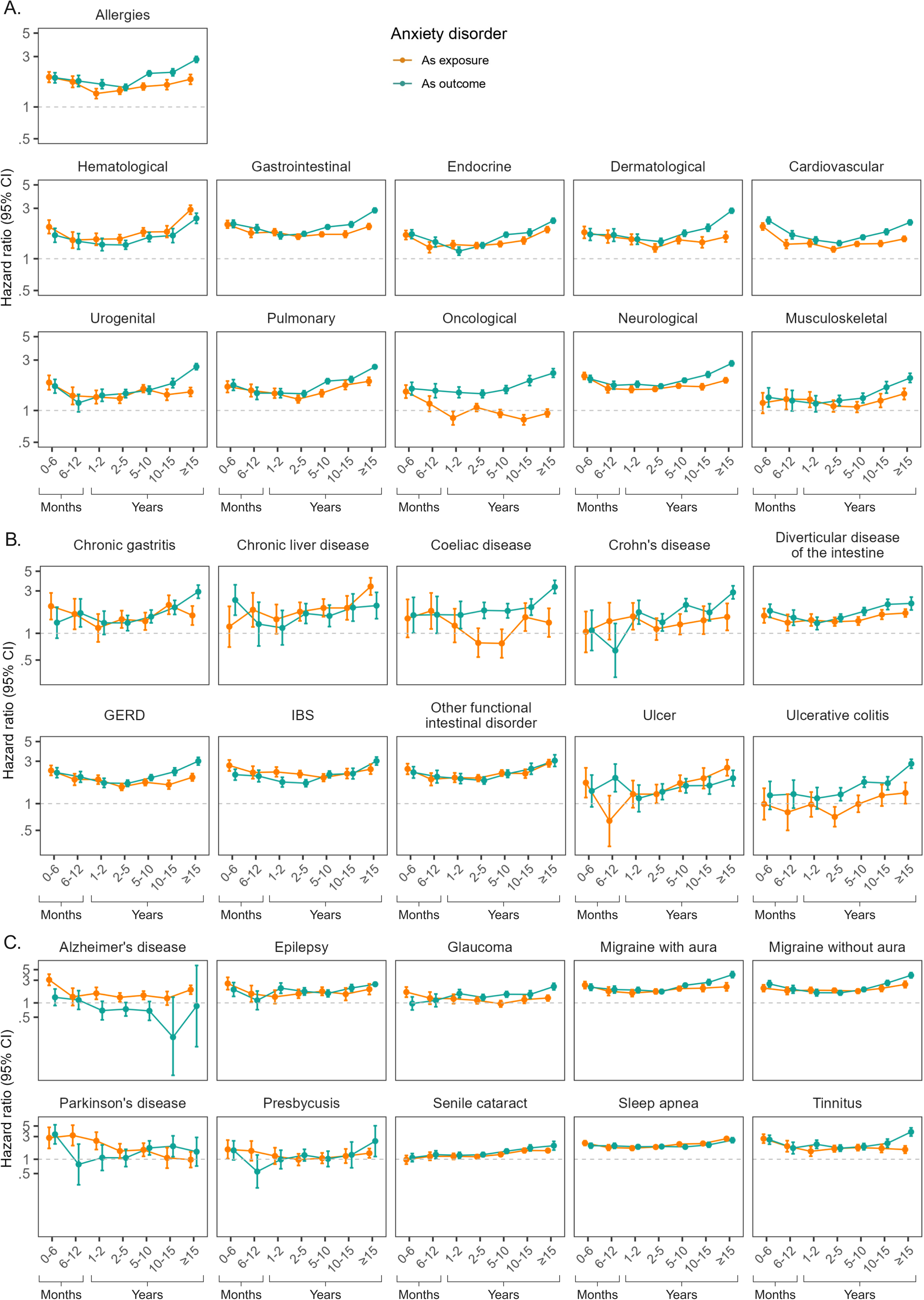
Bidirectional time-dependent Cox proportional hazards analyses. Hazard ratios (HRs) and 95% confidence intervals (CIs) are shown on the y-axis on logarithmic scale and in seven different time intervals on the x-axis representing varying lengths of exposure to the preceding condition. Each panel shows the temporal pattern in HRs bidirectionally; the teal color indicates results from analyses where anxiety disorders are the outcome, and orange line results from analyses where anxiety disorders are the exposure. HRs are estimated by comparing individuals with exposure to the preceding condition to those who are unexposed to that condition. All models are adjusted with sex, calendar time and other broad categorical diagnoses of GMCs and psychiatric conditions with onset before the exposure (model B, see eTables 11-12 for results of the other models) and use age as the underlying timescale. Line of unity is shown in each panel with dotted lines. **A.** Results from the broad category level analyses, **B.** results of individual gastrointestinal diseases and **C.** results of individual neurological diseases. IBS: Irritable bowel syndrome, GERD: gastro-esophageal reflux. Created with BioRender.com.

We further examined the temporal patterns in associations within all 47 individual diseases (Figure 3B-C, eTable 12, eFigures 10-18). In gastrointestinal diseases, the most consistently elevated HRs across time were observed bidirectionally between IBS, other functional intestinal disorder or GERD and anxiety disorders (Figure 3B, eTable 12). In neurological diseases, the strongest and most consistently elevated associations were observed with prior anxiety disorders and later migraine with aura, migraine without aura, and tinnitus (Figure 3C, eTable 12). All HRs specified here were statistically significant with a P < 2x10^-16^.

### Shared Genetic Risk Factors

We observed positive genetic correlation (r_g_ > 0.40, P < 2x10^-5^) between anxiety disorders and some gastrointestinal diseases, including other functional intestinal diseases (r_g_ = 0.59-0.73), IBS (r_g_ = 0.61-0.69), ulcer (r_g_ = 0.43-0.50), GERD (r_g_ = 0.41-0.50), and chronic gastritis (r_g_ = 0.41-0.47) (Figure 4, eTable 13). From neurological disorders, we observed a moderate positive genetic correlation (r_g_ > 0.30, P < 2x10^-5^) between anxiety disorders and tinnitus (r_g_ = 0.32-0.60), migraine with aura (r_g_ = 0.34-0.39), and migraine without aura (r_g_ = 0.31-0.37) (Figure 4, eTable 13). A low to moderate positive genetic correlation (r_g_ > 0.2, P < 0.01) was also observed between anxiety disorders and multiple other diseases (Figure 4, eTable 13, eFigure 19).

**Figure 4.**
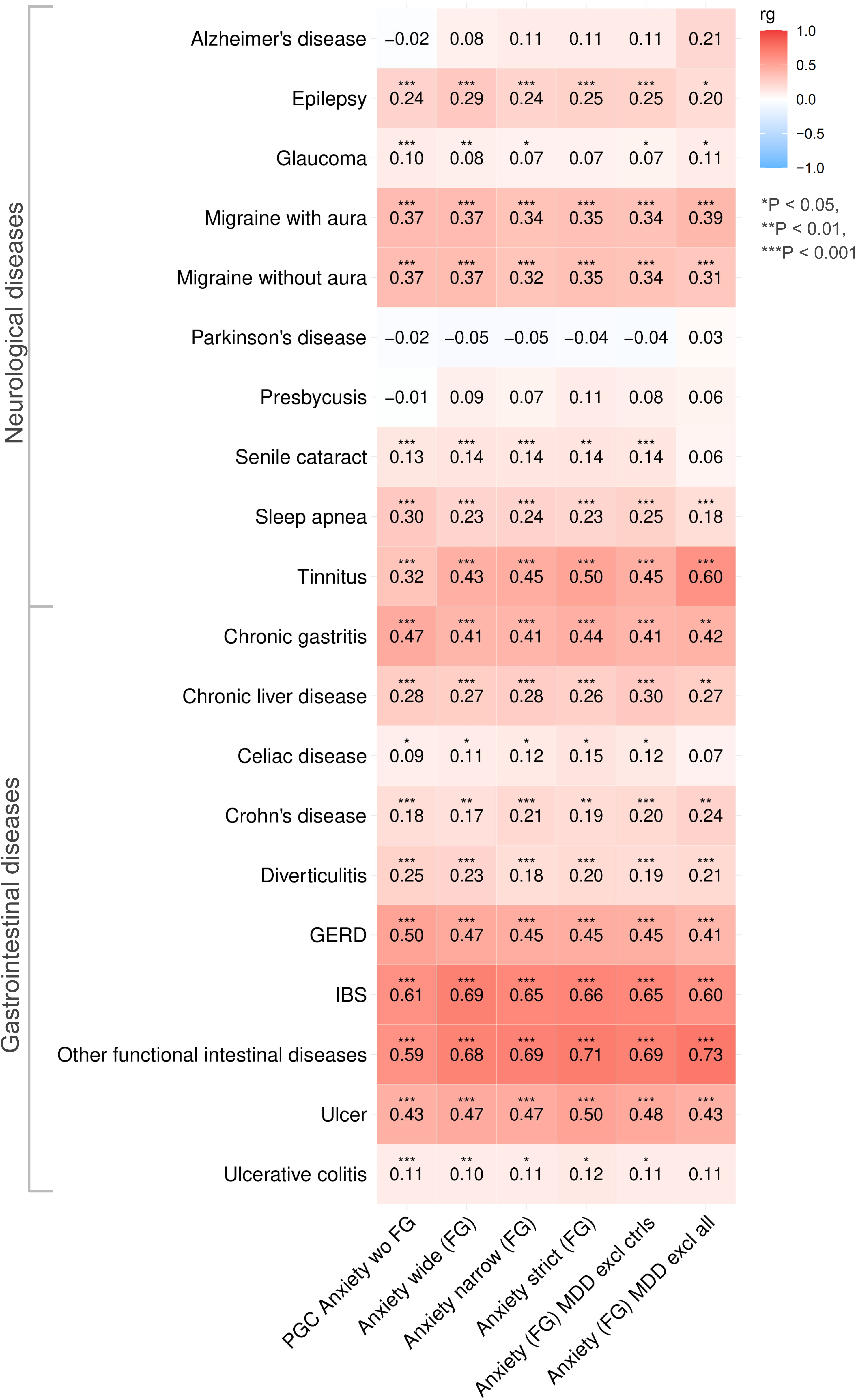
Anxiety disorders have a significant genetic correlation with multiple general medical conditions. Genetic correlation (r_g_) between different anxiety disorder definitions (x-axis) and FinnGen custom neurological and gastrointestinal diseases (y-axis). We used six different anxiety disorder definitions: custom FinnGen GWAS summary-statistics with curated five anxiety disorder phenotypes from hospital and primary care data (Anxiety wide, Anxiety narrow, Anxiety strict, Anxiety with major depressive disorder (MDD) excluded from controls, and Anxiety with MDD excluded from both cases and controls, see eTable 3 for detailed phenotype definitions) and the PGC anxiety disorder meta-analysis leave-one out FinnGen summary statistics^6^. GMC: general medical condition, IBS: Irritable bowel syndrome, GERD: gastro-esophageal reflux, PGC: Psychiatric Genomics Consortium, FG: FinnGen, wo: without, MDD: Major depressive disorder, excl: excluded, ctrls: controls, all: cases and controls. Created with BioRender.com.

Observations of these correlations remained similar in magnitude between the studied traits independent of the anxiety disorder definition used (Figure 4, eTable 13, eFigure 19). For validation of the observed correlations, we analyzed other independent publicly available GWAS summary statistics (eTable 8), and the results aligned with our initial LDSC results particularly with IBS (r_g_ = 0.37-0.64) and GERD (r_g_ = 0.31-0.50) (Figure 5A, eTable 14, eFigure 20).

**Figure 5.**
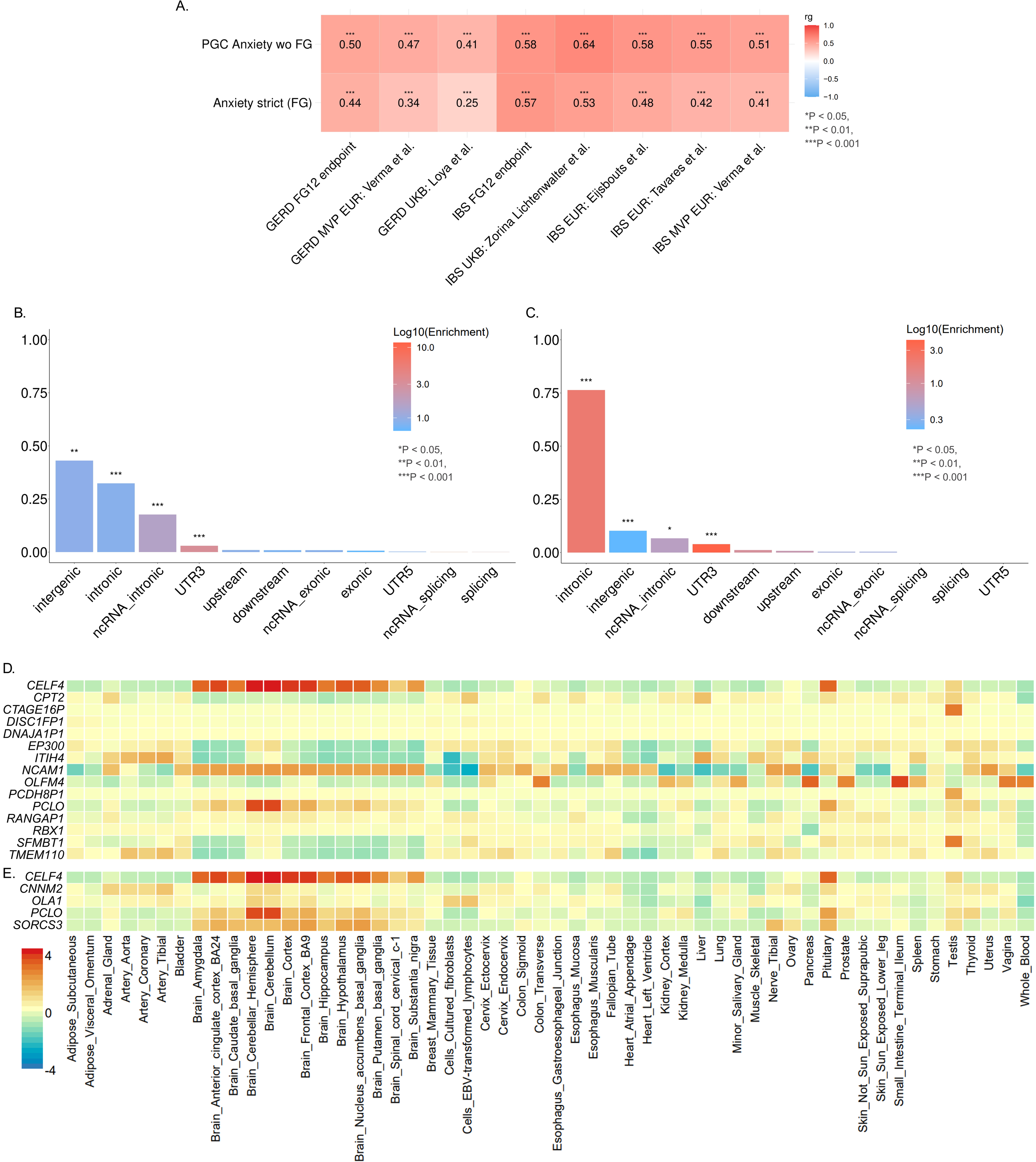
The shared genetics between anxiety disorders and gastrointestinal diseases highlight regulatory regions of the genome. **A.** Genetic correlation (r_g_) between anxiety disorders (y-axis), the PGC anxiety disorder meta-analysis leave-one-out FinnGen ^6^ data and Anxiety strict from the custom FinnGen data, and different GERD or IBS GWAS results from predefined FinnGen endpoints (https://r12.risteys.finngen.fi/) or independent cohorts (x-axis)^18,21,24,25,29^. The proportion (y-axis) and annotation (x-axis) of the shared SNPs between **B.** anxiety disorders and IBS, and **C.** anxiety disorders and GERD. The enrichment (log10) is calculated by dividing proportion of candidate SNPs with the corresponding annotation with the proportion of SNPs with the corresponding annotation in the reference panel (European 1000 Genomes Project Phase 3 Reference Panel^35^). P-value is calculated with two-sided Fisher’s exact test for each annotation against the reference panel. Average of normalized gene expression levels (zero mean across samples, the GTEx project^36^) of the shared genes between **D.** anxiety disorders and IBS, and **E.** anxiety disorders and GERD (eTable 17). Cells with red represent higher expression within a gene across the tissues. IBS: Irritable bowel syndrome, GERD: gastro-esophageal reflux, PGC: Psychiatric Genomics Consortium, FG: FinnGen, wo: without, MVP: Million Veterans Program, EUR: European ancestry, UKB: UK Biobank, ncRNA: Non-coding RNA, UTR: Untranslated region. Created with BioRender.com.

### Regulatory Regions Drive the Genetic Overlap

The highest genetic correlations (r_g_ > 0.3 with two or more independent GMC summary statistics) were witnessed between anxiety disorders and IBS or GERD (Figure 5A, eTable 14, eFigures 19-20). To identify specific variants responsible for this genetic correlation, we conducted pleiotropy analyses from these combinations (eFigures 21-22). We concentrated on the genome-wide significant variants (P < 5x10^-8^) from the pleiotropy analyses that had statistical significance (P < 1x10^-4^) independently in both original GWAS studied (in anxiety disorders or IBS; in anxiety disorders or GERD) (eTables 15,16). This was done to divert from the traits that were significantly driven by one or the other trait (eTables 15,16). Most of the studied shared variants between anxiety disorders and IBS were in the intergenic (43.1 %), intronic (32.4 %), non-coding RNA intronic (17.7 %), or within the 3’ UTR (3.0 %) regions (Figure 5B, eTable 15) as did the shared variants observed between anxiety disorders and GERD (10.2 %, 76.4 %, 6.7 %, and 3.9 %, respectively) (Figure 5C, eTable 16).^35^

We investigated further these genetic variants focusing on those significant genomic loci that had multiple SNPs (> 10) and to genes that mapped on those loci (closest gene by genomic position to corresponding SNP). From the shared genetic loci between anxiety disorders and IBS, 21 genes matched our criteria (STable 15) and out of 15 of them we were able to identify gene expression patterns at a tissue level (GTEx data^36^, Figure 5D, eTable 17, eFigure 23A). Specifically, this tissue level expression was observed in *CELF4*, *NCAM1* and *PCLO* genes and multiple different brain regions such as cerebellar hemisphere (average normalized expression 4.42, 2.39, and 3.88, respectively) and cerebellum (average normalized expression 4.43, 2.44, and 3.91, respectively) (Figure 5D, eTable 17). We, furthermore, observed an enrichment of these genes in positional gene sets chr3p21 (*ITIH4*, *TMEM110-MUSTN1*, *TMEM110,* and *SFMBT1*) and chr22q13 (*RBX1*, *EP300,* and *RANGAP1*). Also, the Biocarta PDZS pathway, which is linked to synaptic function^37^, was mapped to *NCAM1* and *PCLO* genes (P_FDRadj._ = 0.012).

From the shared loci between anxiety disorders and GERD, 6 genes matched our criteria (eTable 16) and 5 of them had gene expression patterns at a tissue level (Figure 5E, eTable 16, eFigure 23B), for example genes *CELF4*, *PCLO,* and *SORCS3* in the pituitary (normalized expression 3.45, 2.24, and 1.81, respectively). Additionally, we identified upregulated differentially expressed genes compared to all other tissues in tissue-specific enrichment analysis for anxiety disorders and GERD in cerebellum (P_FDRadj._ = 0.02), cerebellar hemisphere (P_FDRadj._ = 0.02) and frontal cortex (P_FDRadj._ = 0.03) tissues.

### Causality Estimation

The most consistent causal relationship, regardless of the summary statistics or MR method used, was observed from anxiety disorders to asthma (e.g. from PGC Anxiety without FinnGen to FinnGen asthma: inverse variance weighted beta [SE] = 0.18 [0.05], P_Bonferroni_ = 0.02) (eTables 18-19). In all these associations, the increased risk for anxiety disorders increased the risk for asthma without evidence of pleiotropy (eTables 18-19). We also observed plausible causal relationships from anxiety disorders to diverticulitis, GERD, migraine, and sleep apnea (eTables 18-19). No systematic statistically significant causal associations across different cohorts were observed from GMCs to anxiety disorders (eTables 20-21).

## Discussion

In this broad and systematic study involving > 500,000 individuals from FinnGen, we showed that people with anxiety disorders were more likely to have medical conditions from all other broad disease categories, except cancers, than people without anxiety disorders. We found bidirectional relationships with anxiety disorders, and all investigated broad GMC categories, most notably with gastrointestinal and neurological diseases, regardless of anxiety disorder as exposure or outcome. The time-dependent risk stayed elevated across a time span of over 15 years for most broad disease categories with a prior or later diagnosis of anxiety disorder. The high hazards were partially explained by a significant proportion of shared genetic risk factors between anxiety disorders and some gastrointestinal diseases, particularly IBS and GERD, but also with some neurological disorders, such as migraine with or without aura.

We found consistently the strongest associations as well as genetic correlation between anxiety disorders and gastrointestinal diseases. Interestingly, immune system-related gastrointestinal diseases such as celiac disease and ulcerative colitis increased the hazard for later developing an anxiety disorder, but an anxiety disorder was not associated with the increased risk of subsequently developing these diseases. They also had negligible genetic correlation with anxiety disorders. One reason for the unidirectional association between ulcerative colitis and celiac disease may be that the symptoms of these diseases may lead to central sensitization^38,39^, thereby increasing the hazard for later anxiety disorder. On the contrary, IBS and GERD were associated with higher anxiety disorder risk in the future and anxiety disorder diagnosis was associated with higher risk of IBS and GERD later. Also, one of the highest observed genetic correlations were between anxiety disorders and IBS and between anxiety disorders and GERD, suggesting considerable shared genetic background. Our results agree with recent genetic studies of IBS and GERD in other population cohorts.^9,24,40^ For instance, anxiety was shown to be twice as common among those individuals that had IBS and shared positive genetic correlation (r_g_ = 0.58) with one another in the United Kingdom Biobank sample.^24^

Our pleiotropy analysis between anxiety disorders and IBS or GERD pinpointed genes that were highly expressed in different brain regions. Particularly gene *CELF4*, protein functioning as RNA binding protein in synaptic functions and development^41^, has previously been indicated as an important regulator between depression, neuroticism and gastrointestinal diseases along with genes *PCLO*, *NCAM1* and *SORCS3*.^42^ Also, these proteins encoded by the genes, NCAM1 as a neural cell adhesion molecule, PCLO as a presynaptic protein and SORCS3 as a postsynaptic receptor, are not only localized in synapses but also have a significant role in synaptic functions as well as neural development.^43–45^ One possible mechanism behind the association between anxiety disorders and gastrointestinal diseases may be dysfunction of the gut-brain axis, which has been associated with psychiatric conditions.^46,47^

High hazard and moderate genetic correlation were also witnessed with anxiety disorders and different neurological diseases such as migraine (with or without aura), sleep apnea, and tinnitus aligning with smaller scale studies^48–52^, but less so with neurodegenerative diseases highly influenced by age, such as Alzheimer’s disease and Parkinson’s disease. Additionally, other broad disease categories and individual diseases, such as cardiovascular and hematological diseases, had associations with anxiety disorders emphasizing their multifactorial systemic effects on wellbeing.

## Limitations

The following limitations are important to take into consideration when interpreting our results. Our analyses were mainly conducted using the data from the FinnGen study^11^ involving only data derived from individuals of the European ancestry, and thus, our results are not directly applicable to other populations or ancestries. We acknowledge the known health registries’ limitations in follow-up studies.^53,54^ Finnish individuals suffering from anxiety symptoms tend to contact primary health care services more often compared to the rest of the population providing a plausible bias.^55,56^ The observed SNP-based genetic heritability (for LDSC) as well as the number of valid SNPs as genetic instruments (for MR) may differ depending on the disease studied and the results should be interpreted with this in mind.

## Conclusions

In conclusion, our results indicate a higher somatic disease burden in anxiety disorder patients. By combining longitudinal health record data spanning multiple decades with genomic information, we furthermore identified high genetic correlation between especially some gastrointestinal diseases and anxiety disorders and identified specific underlying genetic variants. Future functional studies will be instrumental in understanding the cellular and molecular mechanisms involved. Overall, our results may help the clinicians identify prevention and treatment needs of anxiety disorder patients.

## Author Contributions

Drs Tervi and Hovatta as well as Suomalainen and Perälampi had full access to all the data in the study and take full responsibility for the integrity of the data and accuracy of the data analysis. Dr Tervi, Suomalainen and Perälampi contributed equally.

Concept and design: Tervi, Suomalainen, Perälampi, Frilander, Suvisaari, Elovainio, Hovatta

Acquisition, analysis, or interpretation of data: Tervi, Suomalainen, Perälampi, Frilander, Naamanka, Suvisaari, Elovainio, Hovatta

Drafting of the manuscript: Tervi, Suomalainen, Perälampi, Hovatta

Critical review of the manuscript for important intellectual content: All authors

Statistical analysis: Tervi, Suomalainen, Perälampi, Frilander

Obtained funding: Hovatta

Administrative, technical, or material support: FinnGen, Anxiety Disorders Working

Group of the Psychiatric Genomics Consortium

Supervision: Hovatta

## Disclaimer

The content is solely the responsibility of the authors.

## Data Sharing Statement

See Supplement 1.

## Data Availability

Based on National and European regulations (GDPR), access to individual-level sensitive health data must be approved by national authorities for specific research projects and for specifically listed and approved researchers. The health data described here was generated and provided by the National Health Register Authorities (Finnish Institute of Health and Welfare, Statistics Finland, KELA, Digital and Population Data Services Agency) and approved, either by the individual authorities or by the Finnish Data Authority, Findata, for use in the FinnGen project. Therefore, we, the authors of this paper, are not in a position to grant access to individual-level data to others. However, any researcher can apply for the health register data from the Finnish Data Authority Findata (https://findata.fi/en/permits/) and for individual-level genotype data from Finnish biobanks via the Fingenious portal (https://site.fingenious.fi/en/) hosted by the Finnish Biobank Cooperative FINBB (https://finbb.fi/en/). All Finnish biobanks can provide access for research projects within the scope regulated by the Finnish Biobank Act, which is research using the biobank samples or data for the purposes of promoting health, understanding the mechanisms of disease, or developing products and treatment practices used in health and medical care. You can learn more about accessing other FinnGen data here: https://www.finngen.fi/en/access_results.
Summary-level data will be deposited in the FinnGen public bucket and will be publicly available as of the date of publication. The code used in this study will be available upon publication in a GitHub repository.

## Acknowledgements

We thank Dr. Samuel E. Jones for his valuable advise conducting the genetic analyses and Hovatta lab members for helpful discussions. We thank the Anxiety Disorders Working Group of the Psychiatric Genomics Consortium (eTable 22) for summary level data sharing.

We acknowledge the participants and investigators of the FinnGen study (eTable 23). The FinnGen project is funded by two grants from Business Finland (HUS 4685/31/2016 and UH 4386/31/2016) and the following industry partners: AbbVie Inc., AstraZeneca UK Ltd, Biogen MA Inc., Bristol Myers Squibb (and Celgene Corporation & Celgene International II Sàrl), Genentech Inc., Merck Sharp & Dohme LCC, Pfizer Inc., GlaxoSmithKline Intellectual Property Development Ltd., Sanofi US Services Inc., Maze Therapeutics Inc., Janssen Biotech Inc, Novartis AG, and Boehringer Ingelheim International GmbH. Following biobanks are acknowledged for delivering biobank samples to FinnGen: Auria Biobank (www.auria.fi/biopankki), THL Biobank (www.thl.fi/biobank), Helsinki Biobank (www.helsinginbiopankki.fi), Biobank Borealis of Northern Finland (https://www.ppshp.fi/Tutkimus-ja-opetus/Biopankki/Pages/Biobank-Borealis-briefly-in-English.aspx), Finnish Clinical Biobank Tampere (www.tays.fi/en-US/Research_and_development/Finnish_Clinical_Biobank_Tampere), Biobank of Eastern Finland (www.ita-suomenbiopankki.fi/en), Central Finland Biobank (www.ksshp.fi/fi-FI/Potilaalle/Biopankki), Finnish Red Cross Blood Service Biobank (www.veripalvelu.fi/verenluovutus/biopankkitoiminta), Terveystalo Biobank (www.terveystalo.com/fi/Yritystietoa/Terveystalo-Biopankki/Biopankki/) and Arctic Biobank (https://www.oulu.fi/en/university/faculties-and-units/faculty-medicine/northern-finland-birth-cohorts-and-arctic-biobank). All Finnish Biobanks are members of BBMRI.fi infrastructure (https://www.bbmri-eric.eu/national-nodes/finland/). Finnish Biobank Cooperative -FINBB (https://finbb.fi/) is the coordinator of BBMRI-ERIC operations in Finland. The Finnish biobank data can be accessed through the Fingenious® services (https://site.fingenious.fi/en/) managed by FINBB.

