## Supplementary_information for "Bidirectional Association and Genetic Background of Anxiety Disorders and General Medical Conditions"

### Supplementary Material

eMethods ... 1
Sample ... 1
Ethics statement ... 1
Phenotypes ... 2
Epidemiological analyses ... 2
Genetic Analyses ... 3
Data and Code Availability ... 5
eFigures ... 6
eFigure 1. Cox regression: Broad GMC categories ... 6
eFigure 2. Cox regression: Gastrointestinal diseases ... 7
eFigure 3. Cox regression: Neurological diseases ... 8
eFigure 4. Cox regression: A. Cardiovascular and B. endocrinological diseases ... 9
eFigure 5. Cox regression: A. Pulmonary, B. urogenital and C. hematological diseases ... 10
eFigure 6. Cox regression: A. Musculoskeletal and B. dermatological diseases ... 11
eFigure 7. Bidirectional time-dependent Cox regression: Broad GMC categories (model A) ... 12
eFigure 8. Bidirectional time-dependent Cox regression: Broad GMC categories (model C) ... 13
eFigure 9. Bidirectional time-dependent Cox regression: Broad GMC categories (model D) ... 13
eFigure 10. Bidirectional time-dependent Cox regression: Gastrointestinal diseases (models A, C and D) ... 15
eFigure 11. Bidirectional time-dependent Cox regression: Neurological diseases (models A, C and D) ... 16
eFigure 12. Bidirectional time-dependent Cox regression: Cardiovascular diseases (all models) ... 17
eFigure 13. Bidirectional time-dependent Cox regression: Endocrine diseases (all models) ... 18
eFigure 14. Bidirectional time-dependent Cox regression: Pulmonary diseases (all models) ... 19
eFigure 15. Bidirectional time-dependent Cox regression: Urogenital diseases (all models) ... 20
eFigure 16. Bidirectional time-dependent Cox regression: Musculoskeletal diseases (all models) ... 21
eFigure 17. Bidirectional time-dependent Cox regression: Hematological diseases (all models) ... 22
eFigure 18. Bidirectional time-dependent Cox regression: Dermatological diseases (all models) ... 23
eFigure 19. Initial genetic correlation results ... 24
eFigure 20. Genetic correlation with publicly available data and FinnGen endpoints ... 25
eFigure 21. CPASSOC PGC Anxiety and meta-analysis of IBS ... 26
eFigure 22. CPASSOC PGC Anxiety and meta-analysis of GERD ... 27
eFigure 23. Tissue-level gene expression ... 28
References ... 29

Bidirectional association and genetic background of anxiety disorders and general medical conditions
Tervi *et al*.

#### eMethods

##### Sample

The FinnGen[1] initiative is a large-scale public-private partnership launched in Finland in 2017. It combines genome-wide data with longitudinal electronic health record (EHR) data, including for instance primary health care, hospital, death, cancer and drug reimbursement registries, from approximately 500,000 Finnish participants representing almost 10 % of the Finnish population. The registries in the FinnGen study are comprehensive due to the centralized nature of the Finnish health care system where the health information of individuals is linked to their unique social security number throughout their lifespan. The consortium includes Finnish universities, university hospitals, biobanks, the Finnish Biobank Cooperative (FINBB), and international pharmaceutical companies (a complete list of partners is available at https://www.finngen.fi/en/partners).

##### Ethics statement

Study subjects in FinnGen provided informed consent for biobank research, based on the Finnish Biobank Act. Alternatively, separate research cohorts, collected prior the Finnish Biobank Act came into effect (in September 2013) and start of FinnGen (August 2017), were collected based on study-specific consents and later transferred to the Finnish biobanks after approval by Fimea (Finnish Medicines Agency), the National Supervisory Authority for Welfare and Health. Recruitment protocols followed the biobank protocols approved by Fimea. The Coordinating Ethics Committee of the Hospital District of Helsinki and Uusimaa (HUS) statement number for the FinnGen study is Nr HUS/990/2017.

The FinnGen study is approved by Finnish Institute for Health and Welfare (permit numbers: THL/2031/6.02.00/2017, THL/1101/5.05.00/2017, THL/341/6.02.00/2018, THL/2222/6.02.00/2018, THL/283/6.02.00/2019, THL/1721/5.05.00/2019 and THL/1524/5.05.00/2020), Digital and population data service agency (permit numbers: VRK43431/2017-3, VRK/6909/2018-3, VRK/4415/2019-3), the Social Insurance Institution (permit numbers: KELA 58/522/2017, KELA 131/522/2018, KELA 70/522/2019, KELA 98/522/2019, KELA 134/522/2019, KELA 138/522/2019, KELA 2/522/2020, KELA 16/522/2020), Findata permit numbers THL/2364/14.02/2020, THL/4055/14.06.00/2020, THL/3433/14.06.00/2020, THL/4432/14.06/2020, THL/5189/14.06/2020, THL/5894/14.06.00/2020, THL/6619/14.06.00/2020, THL/209/14.06.00/2021, THL/688/14.06.00/2021, THL/1284/14.06.00/2021, THL/1965/14.06.00/2021, THL/5546/14.02.00/2020, THL/2658/14.06.00/2021, THL/4235/14.06.00/2021, Statistics Finland (permit numbers: TK-53-1041-17 and TK/143/07.03.00/2020 (earlier TK-53-90-20) TK/1735/07.03.00/2021, TK/3112/07.03.00/2021) and Finnish Registry for Kidney Diseases.

The Biobank Access Decisions for FinnGen samples and data utilized in FinnGen Data Freeze 12 include: THL Biobank BB2017_55, BB2017_111, BB2018_19, BB_2018_34, BB_2018_67, BB2018_71, BB2019_7, BB2019_8, BB2019_26, BB2020_1, BB2021_65, Finnish Red Cross Blood Service Biobank 7.12.2017, Helsinki Biobank HUS/359/2017, HUS/248/2020, HUS/430/2021 §28, §29, HUS/150/2022 §12, §13, §14, §15, §16, §17, §18, §23, §58, §59, HUS/128/2023 §18, Auria Biobank AB17-5154 and amendment #1 (August 17 2020) and amendments BB_2021-0140, BB_2021-0156 (August 26 2021, Feb 2 2022), BB_2021-0169, BB_2021-0179, BB_2021-0161, AB20-5926 and amendment #1 (April 23 2020) and it´s modifications (Sep 22 2021), BB_2022-0262, BB_2022-0256, Biobank Borealis of Northern Finland_2017_1013, 2021_5010, 2021_5010 Amendment, 2021_5018, 2021_5018 Amendment, 2021_5015, 2021_5015 Amendment, 2021_5015 Amendment_2, 2021_5023, 2021_5023 Amendment, 2021_5023 Amendment_2, 2021_5017, 2021_5017 Amendment, 2022_6001, 2022_6001 Amendment, 2022_6006 Amendment, 2022_6006 Amendment, 2022_6006 Amendment_2, BB22-0067, 2022_0262, 2022_0262 Amendment, Biobank of Eastern Finland 1186/2018 and amendment 22§/2020, 53§/2021, 13§/2022, 14§/2022, 15§/2022, 27§/2022, 28§/2022, 29§/2022, 33§/2022, 35§/2022, 36§/2022, 37§/2022, 39§/2022, 7§/2023, 32§/2023, 33§/2023, 34§/2023, 35§/2023, 36§/2023, 37§/2023, 38§/2023, 39§/2023, 40§/2023, 41§/2023, Finnish Clinical Biobank Tampere MH0004 and amendments (21.02.2020 & 06.10.2020), BB2021-0140 8§/2021, 9§/2021, §9/2022, §10/2022, §12/2022,

13§/2022, §20/2022, §21/2022, §22/2022, §23/2022, 28§/2022, 29§/2022, 30§/2022, 31§/2022, 32§/2022, 38§/2022, 40§/2022, 42§/2022, 1§/2023, Central Finland Biobank 1-2017, BB_2021-0161, BB_2021-0169, BB_2021-0179, BB_2021-0170, BB_2022-0256, BB_2022-0262, BB22-0067, Decision allowing to continue data processing until 31st Aug 2024 for projects: BB_2021-0179, BB22-0067,BB_2022-0262, BB_2021-0170, BB_2021-0164, BB_2021-0161, and BB_2021-0169, and Terveystalo Biobank STB 2018001 and amendment 25th Aug 2020, Finnish Hematological Registry and Clinical Biobank decision 18th June 2021, Arctic biobank P0844: ARC_2021_1001.

#### Phenotypes

We defined our phenotypes in FinnGen data release 12 based on International Classification of Diseases (ICD) eighth, ninth and tenth revisions, and the Social Insurance Institution of Finland (Kansaneläkelaitos, KELA) reimbursement codes for healthcare services and medications. We utilized the primary health care, inpatient, outpatient, death, cancer and drug purchase registries. Anxiety disorders were considered as one category, including phobic anxiety disorders (F40) and other anxiety disorders (F41) from ICD-10 (eTable 1). We concentrated on common anxiety disorders affecting adults and excluded disorders mainly manifesting in childhood (separation anxiety disorder and selective mutism) or disorders that are caused by substance or medication use, or by another medical condition (eTable 2). Also, obsessive compulsive disorder (OCD), post-traumatic stress disorder (PTSD), and other neurotic, stress-related and somatoform disorders were not included, to specifically focus on core anxiety disorders. Anxiety disorders were identified across all ICD manuals used in Finland and correspond to the following codes and their subcodes: ICD-10 diagnoses F40 or F41; ICD-9 diagnoses 3000A, 3000B, 3000C, 3002B, 3002D, or 3002X; or ICD-8 diagnoses 3000 or 3002.

Diagnostic codes for GMCs were selected based on the chapters and blocks of ICD codes and prior research[2]. These broad categories consisted of cardiovascular, endocrine, pulmonary, gastrointestinal, urogenital, musculoskeletal, hematological, oncological, neurological and dermatological diseases, and allergies. The codes and broad disease categories were then modified to align with the Finnish specific ICD versions and KELA codes and the prevalence (> 1%) of the disorders in the FinnGen dataset (eTable 1). Individuals diagnosed with autism, intellectual disability, psychosis, or schizophrenia were excluded due to FinnGen's disease-specific samples of these disorders (eTable 2). Somatoform disorders were also excluded because the focus of this study was to identify genuine co-occurring GMCs that give rise to experiences of anxiety, and such cases could obscure the signal we aimed to uncover. All inclusion codes, as well as the number of people diagnosed with these conditions before follow-up, during follow-up, and the number of individuals at risk at the start of follow-up, are presented in eTable 1.

For genetic analyses, the same ICD- and KELA-codes, without exclusions for GMCs, were used to define the phenotypes of interest (eTable 1,2). Individuals were included as cases for GMCs in genetic analyses if they had the diagnosis codes in question, regardless of the date of diagnosis, otherwise the individual was included as a control. For different anxiety disorder definitions in FinnGen, the cases and controls were defined with specific inclusion and exclusion criteria (for further information see eTable 3). Final numbers of cases and controls for genetic analyses can be found in eTable 4. Additionally, we used the Psychiatric Genomics Consortium (PGC) Anxiety Disorder Working Group (PGC-ANX) meta-analysis leave-one out FinnGen[3] summary statistics as well as publicly available GMC summary statistics (obtained from the GWAS (genome-wide association study) catalog https://www.ebi.ac.uk/gwas/home), and FinnGen defined endpoints (for further information see https://r12.risteys.finngen.fi/) (eTables 5,6).

#### Epidemiological analyses

##### *Demographics and disease prevalence*

Initially, we examined the prevalence of 11 ICD-code based broad diagnostic categories corresponding to different GMCs (eTable 1) among individuals diagnosed with an anxiety disorder. Within each diagnostic broad category, the individually defined phenotypes were pooled, meaning that each participant was counted only once as having a categorical diagnosis, based on whichever individual disease was diagnosed first. The proportion of GMC diagnoses among cases with anxiety disorders was compared with that of twice the number of age (+/- 2 years) and sex matched controls using the Wilson score interval[4]. Statistical significance ($P < 0.05$) of differences in proportions was tested using the Chi-squared test (Figure 1, eTable 7). All analyses were conducted using R (version 4.5.1).

##### *Cox proportional hazards models*

Cox proportional hazards models[5], with age as the underlying time scale, were employed to assess the bidirectional associations between anxiety disorders and the GMCs of interest. In the broad categories, the follow-up time spanned 12 years, from 01/01/2011 until 12/31/2022. The start of the follow-up time was defined based on having the full information available for all the registries used in our study within FinnGen.

Minimum age thresholds for left truncation were defined based on age-of-onset distributions among cases diagnosed during follow-up. If the two youngest cases were diagnosed within one year of each other and the youngest was < 1 years old when diagnosed, the threshold was set to 0. Otherwise, the threshold was set to the age of the youngest or second youngest case, depending on whether the time difference between them exceeded one year. These thresholds, rounded down to full years, determined when individuals could enter the follow-up period, either on 01/01/2011 or upon reaching the approximated diagnosis-specific minimum age.

All models were adjusted for date of entry to the follow-up period and sex. In addition, the analyses were adjusted further with preceding categorical comorbidities and the number of these comorbid GMCs and psychiatric conditions (2, 3 or 4+). In total, four different models were created: (A) unadjusted (no further adjustments with comorbidities), (B) adjusted for both GMCs and psychiatric comorbidities, (C) adjusted for GMC comorbidities only and (D) adjusted for psychiatric comorbidities only (eTable 8). The outcome variable was included in the model during follow-up. Otherwise, individuals were excluded as prevalent cases. Data from previous conditions were utilized whenever available, and individuals with prior disorders occurring before January 1, 2011, were considered exposed in the model from that date onwards. Adjustment information was collected from the beginning of the registers but was not updated after the occurrence of a prior disorder. The availability of register information varied, with the earliest being the cancer register, which started in 1953, and the most recent being the primary health care register, which began in 2011.

In addition, we conducted bidirectional, time-dependent Cox proportional hazards models to assess the potential temporal effects in hazard ratios (HRs) associated with varying lengths of exposure to the preceding condition. The exposure period was divided into seven intervals, ranging from 0–6 months to ≥ 15 years. HRs were evaluated separately for each exposure interval. We utilized the code provided by Plana-Ripoll et al.[6] in their study. All analyses were conducted using R version 4.5.1 (packages: survival[7], lubridate[8], tidyr[9], data.table[10], reshape2[11], dplyr[12], multcomp[13], stringr[14], MatchIt[15] and binom[16]). Figures for Cox proportional hazard models were made using GraphPad Prism version 10.1.2 for Windows, and the figures for the time-dependent Cox proportional hazard models were configurated with ggplot2[17] R package.

#### Genetic Analyses

##### *Genotyping*

Genotyping in the FinnGen cohort was performed by using Affymetrix (Thermo Fisher Scientific, Santa Clara, CA, USA) and Illumina (Illumina Inc., San Diego, CA, USA) arrays and lifted over to Genome Reference Consortium Human Build version 38 (GRCh38/hg38). For imputation, individuals with excess heterozygosity (+-3 standard deviations), high genotype absence (> 2%), inexplicit sex or duplicates were excluded from the data. Genetic variants with high absence (> 2%), low minor allele count (< 3), low variant-wise call-rate (< 90 %) or low Hardy-Weinberg Equilibrium (HWE) ($P < 1x10^{-9}$) were excluded. All individuals were Finns matched against the SiSu v4 reference panel (http://www.sisuproject.fi/) and the imputation was conducted using Beagle 4.1.[18] For more information see Kurki et al. 2023.[1]

##### *GWAS*

GWAS were conducted using the REGENIE (v.3.3) pipeline for data freeze 12 (R12) FinnGen data (https://github.com/FINNGEN/regenie-pipelines). Analyses were adjusted for age at death or end of follow up (04/28/2023), sex, genotyping batches and the first 10 genetic principal components. Additive model and firth approximation was applied for variants with association P-value < 0.01.[19] These GWAS were run to include the primary care data alongside hospital registries in FinnGen and to reflect better the definitions used in our Cox regression analyses, which partially differ from already publicly available summary statistics conducted by the FinnGen core team (https://www.finngen.fi/en/access_results).

##### *Genetic correlation*

Genetic correlation analyses between anxiety disorders and GMCs were performed using the linkage disequilibrium score regression (LDSC) method provided by the Broad Institute of Massachusetts Institute of Technology (MIT) and Harvard and MRC Integrative Epidemiology Unit, University of Bristol.[20] LDSC method uses the LD structure of the genome to explore the SNP-based heritability of a trait and the possible genetic correlation between two separate traits. For anxiety disorders, we used custom FinnGen R12 GWAS summary-statistics with curated five anxiety disorder phenotypes from hospital and primary care data (ANXIETY_WIDE, ANXIETY_NARROW, ANXIETY_STRICT, ANXIETY_NARROW_MDD_EXCL_CTRL and ANXIETY_NARROW_MDD_EXCL_ALL, see eTable 3 for detailed phenotype definitions) and the PGC-ANX meta-analysis leave-one out FinnGen summary statistics [3]. Initial analyses were conducted using summary statistics from custom FinnGen R12 GWAS and curated GMC phenotypes from hospital, primary care and drug purchase data (see eTable 1 for detailed phenotype definitions). Sensitivity analyses were conducted using publicly available summary statistics[21–33] and FinnGen predefined endpoints for different GMCs (see eTables 5 and 6 for detailed information). Summary statistics from other cohorts for sensitivity analyses were obtained for GMCs that reached $r_g > 0.3$ in the initial LDSC analysis (eTable 7). Additionally, we used custom FinnGen R12 GWAS summary statistics of psychiatric disorders (eTables 2,4) for LDSC between anxiety disorders and psychiatric disorders as a sensitivity analysis to investigate the known correlation between anxiety disorders and psychiatric disorders[34] within the FinnGen sample (eTable 13). HapMap 3 SNP list and European LD score files provided by the software were used in our genetic correlation analyses. Figures for the LDSC result were conducted with R (R v. 4.4.3) using the ggplot2[17] package.

##### *Mendelian Randomization*

For causality estimation we used Mendelian Randomization (MR) and obtained the lead SNPs ($P < 5x10^{-8}$) associated with either anxiety disorders or GMCs. Anxiety disorder SNPs were obtained from the PGC-ANX meta-analysis leave-one out FinnGen [3] or FinnGen custom anxiety definitions (eTable 3). GMC SNPs were obtained from the custom FinnGen R12 GWAS analyses (eTables 1,4) or publicly available summary statistics (eTable 5). Analyses were conducted either using anxiety disorders as exposure or as an outcome as were for the GMCs of interest. Initial MR was conducted for all the individual GMCs utilizing the TwoSampleMR[35,36] R package (R v. 4.4.3). For validation, those traits that had suggested causal effect (inverse variance weighted (IVW) $P_{Bonferroni} < 0.05$) were tested again using publicly available summary statistics (eTable 5). Possible horizontal pleiotropic effects were tested using the Egger intercept methods as part of the R package.

##### *Pleiotropy analyses*

For further investigation of the genetic correlation between anxiety disorders and GMCs, we conducted pleiotropy analyses with the CPASSOC[37] method, which compares the Z-scores of each SNP from GWAS summary statistics taking also into account the possible sample overlap between the traits of interest. CPASSOC has two settings, SHom (Homogenous effective model) and SHet (Heterogeneous effective model), that serve as complementary tools for one another. SHom assumes a consistent and similar direction effect across the phenotypes investigated and is analogous to a fixed-effects meta-analysis[38]. SHet allows more variation in the genetic effects across the phenotypes studied and is more comparable to random effects[38] meta-analysis method. When both settings yielded results the pleiotropic effects are said to be consistent and when only SHet is significant the pleiotropic effects are said to be directional. We ran both settings separately for each phenotype combination of interest, anxiety disorders and irritable bowel syndrome (IBS) as well as anxiety disorders and gastroesophageal reflux disease (GERD) with similar phenotype definitions and $r_g$ > 0.3 in separate LDSC analyses (two or more) in independent samples (eTables 13,14).

Given the multiple sources for GWAS summary statistics for different GMCs, we conducted separate meta-analyses for the traits of interest, IBS (combining three GWAS summary statistics, FinnGen, mixed European meta-analysis[28] and the Million Veteran Program (MVP)[25] with total N of 1,375,771) and GERD (combining three GWAS summary statistics, FinnGen, United Kingdom Biobank[22] and MVP[25] with total N of 1,307,702) specifically, hence, increasing the power to detect disease specific genetic associations and utilize those in our pleiotropy analyses. Effect estimate based meta-analyses were conducted using METAL[39] with standard settings, fixed effect model, and tracking the joint allele frequency with the AVERAGEFREQ option. Heterogeneity between used summary statistics was analyzed with the ANALYZE HETEROGENEITY option. Individual rsIDs were used to match different summary statistics within phenotypes and the final meta-analysis summary statistics were in GRCh38 prior to pleiotropy analyses. The results from pleiotropy analyses were visualized using the qqman[40] R package (R v. 4.4.3) and FUMAGWAS with SNP2GENE and GENE2FUNC[41] utilizing the Genotype-Tissue Expression (GTEx) data[42] for tissue level gene expression analyses.

**Data and Code Availability**

Based on National and European regulations (GDPR), access to individual-level sensitive health data must be approved by national authorities for specific research projects and for specifically listed and approved researchers. The health data described here was generated and provided by the National Health Register Authorities (Finnish Institute of Health and Welfare, Statistics Finland, KELA, Digital and Population Data Services Agency) and approved, either by the individual authorities or by the Finnish Data Authority, Findata, for use in the FinnGen project. Therefore, we, the authors of this paper, are not in a position to grant access to individual-level data to others. However, any researcher can apply for the health register data from the Finnish Data Authority Findata (https://findata.fi/en/permits/) and for individual-level genotype data from Finnish biobanks via the Fingenious portal (https://site.fingenious.fi/en/) hosted by the Finnish Biobank Cooperative FINBB (https://finbb.fi/en/). All Finnish biobanks can provide access for research projects within the scope regulated by the Finnish Biobank Act, which is research using the biobank samples or data for the purposes of promoting health, understanding the mechanisms of disease, or developing products and treatment practices used in health and medical care. You can learn more about accessing other FinnGen data here: https://www.finngen.fi/en/access_results.

Summary-level data will be deposited in the FinnGen public bucket and will be publicly available as of the date of publication. The code used in this study will be available upon publication in a GitHub repository.

### eFigures

#### eFigure 1. Cox regression: Broad GMC categories

Cox proportional hazards models between anxiety disorders and broad general medical condition (GMC) categories. Orange colors depicting anxiety disorders as the exposure (left) and teal colors anxiety disorders as the outcome (right). GMCs are listed on the y-axis and hazard ratios with 95% confidence intervals (CIs) on the x-axis. Models A are adjusted for sex and the date of entry into follow-up, and models B are adjusted for sex, the date of entry into follow-up, and other somatic and psychiatric disorder categories, as well as the number of medical condition categories (2, 3, or 4+). Models C are adjusted for sex, the date of entry into follow-up, all other general medical condition categories, and the number of general medical condition categories (2, 3, or 4+). Models D are adjusted for sex, the date of entry into follow-up, psychiatric disorder categories the number of psychiatric condition categories (2, 3, or 4+).

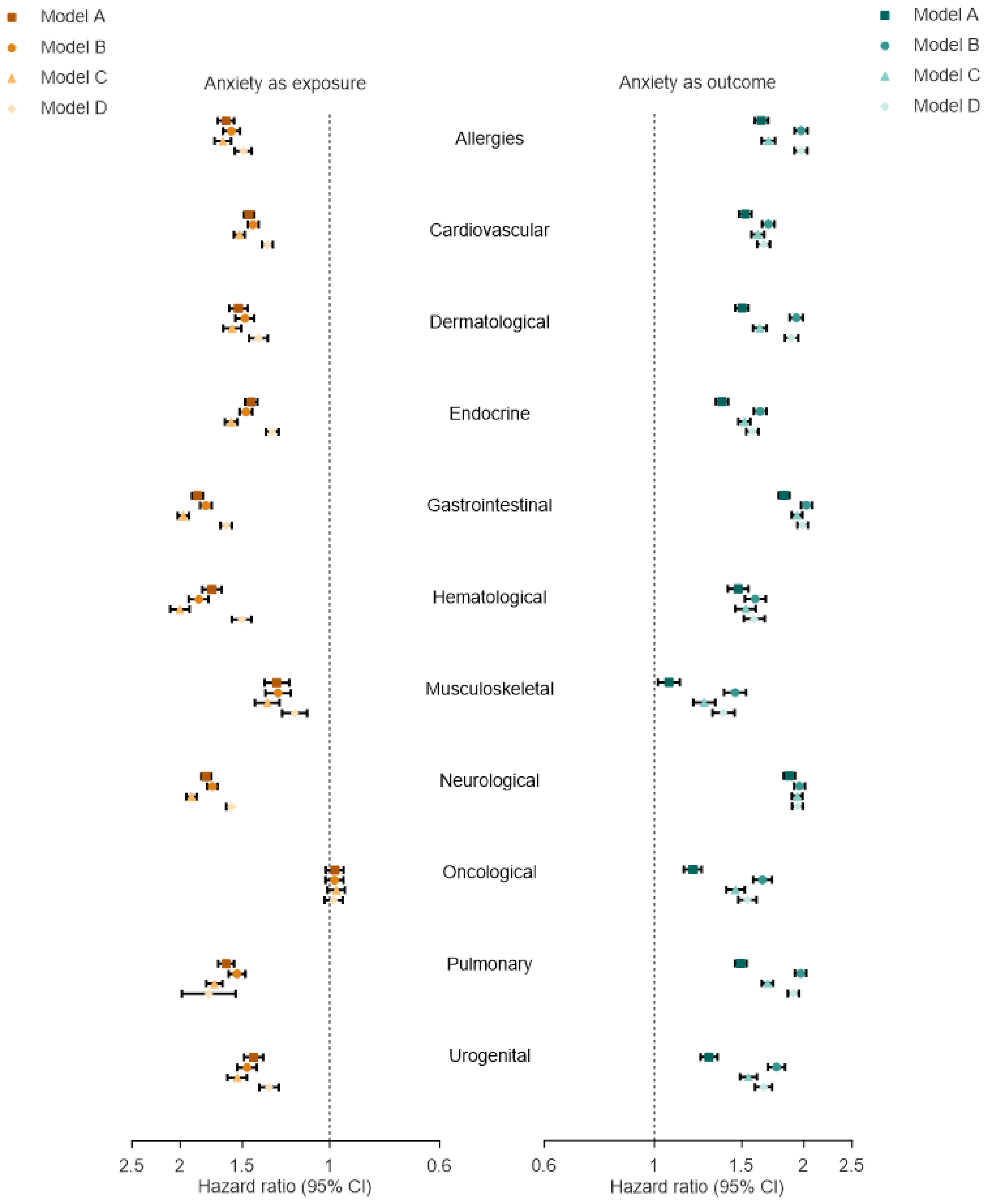

eFigures 2-6 show Cox proportional hazards models between anxiety disorders and individual diseases in different general medical condition categories (eTable 1). Orange colors depicting anxiety disorders as the exposure (left) and teal colors anxiety disorders as the outcome (right). Individual diseases are listed on the y-axis and hazard ratios with 95% confidence intervals (CIs) on the x-axis. Models A are adjusted for sex and the date of entry into follow-up, and models B are adjusted for sex, the date of entry into follow-up, and other somatic and psychiatric disorder categories, as well as the number of medical condition categories (2, 3, or 4+). Models C are adjusted for sex, the date of entry into follow-up, all other general medical condition categories, and the number of general medical condition categories (2, 3, or 4+). Models D are adjusted for sex, the date of entry into follow-up, psychiatric disorder categories the number of psychiatric condition categories (2, 3, or 4+).

#### eFigure 2. Cox regression: Gastrointestinal diseases

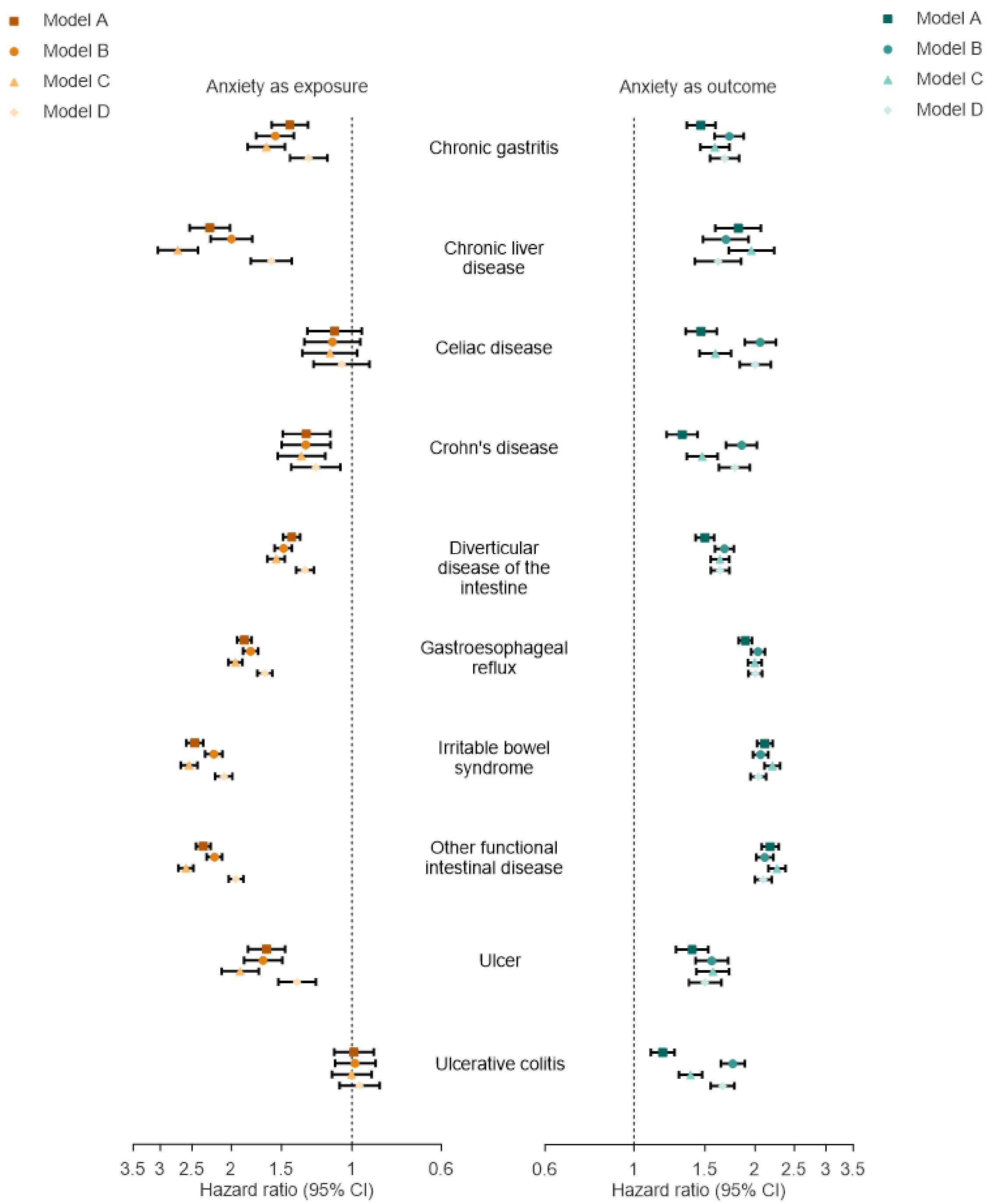

#### eFigure 3. Cox regression: Neurological diseases

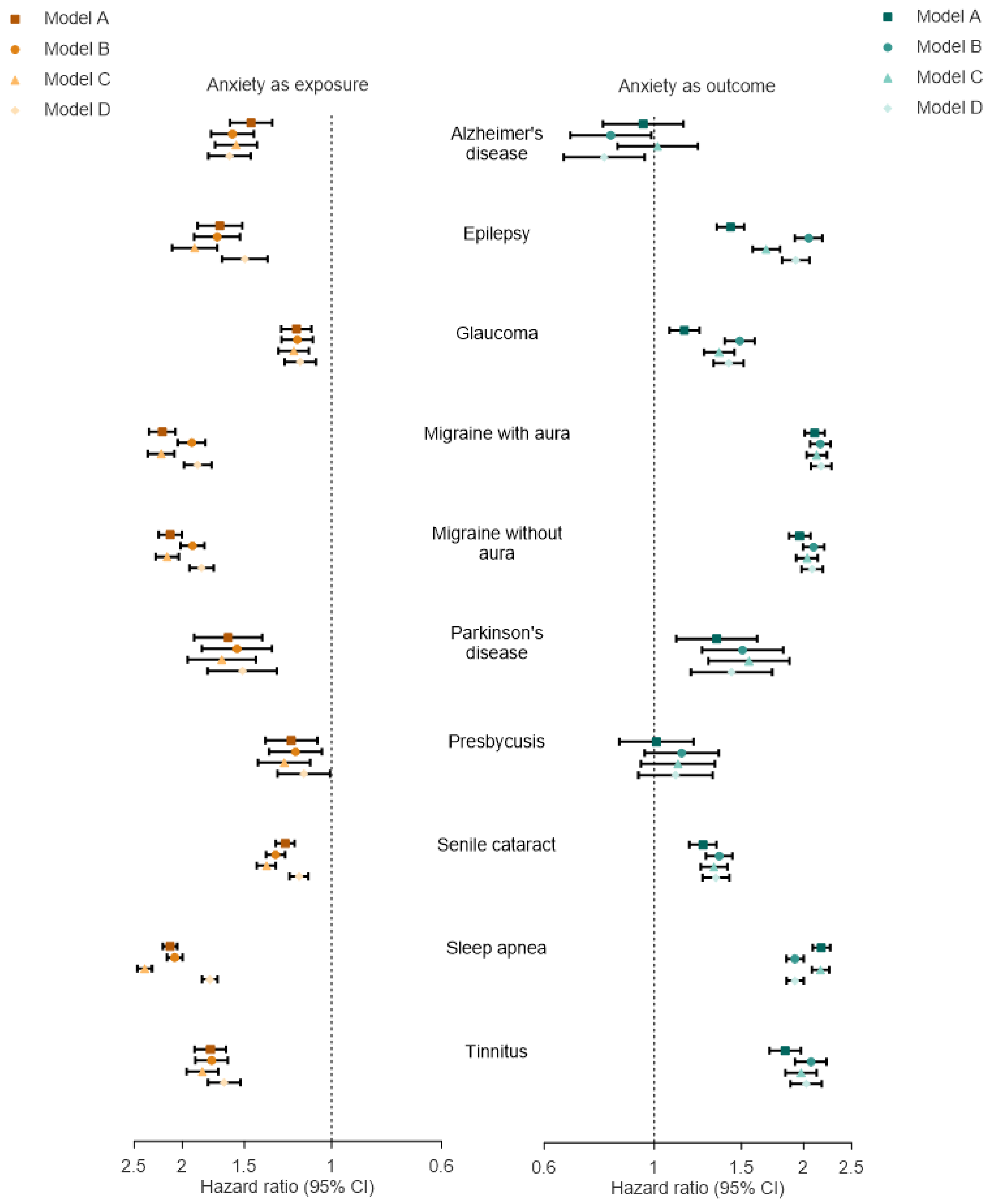

#### eFigure 4. Cox regression: A. Cardiovascular and B. endocrinological diseases

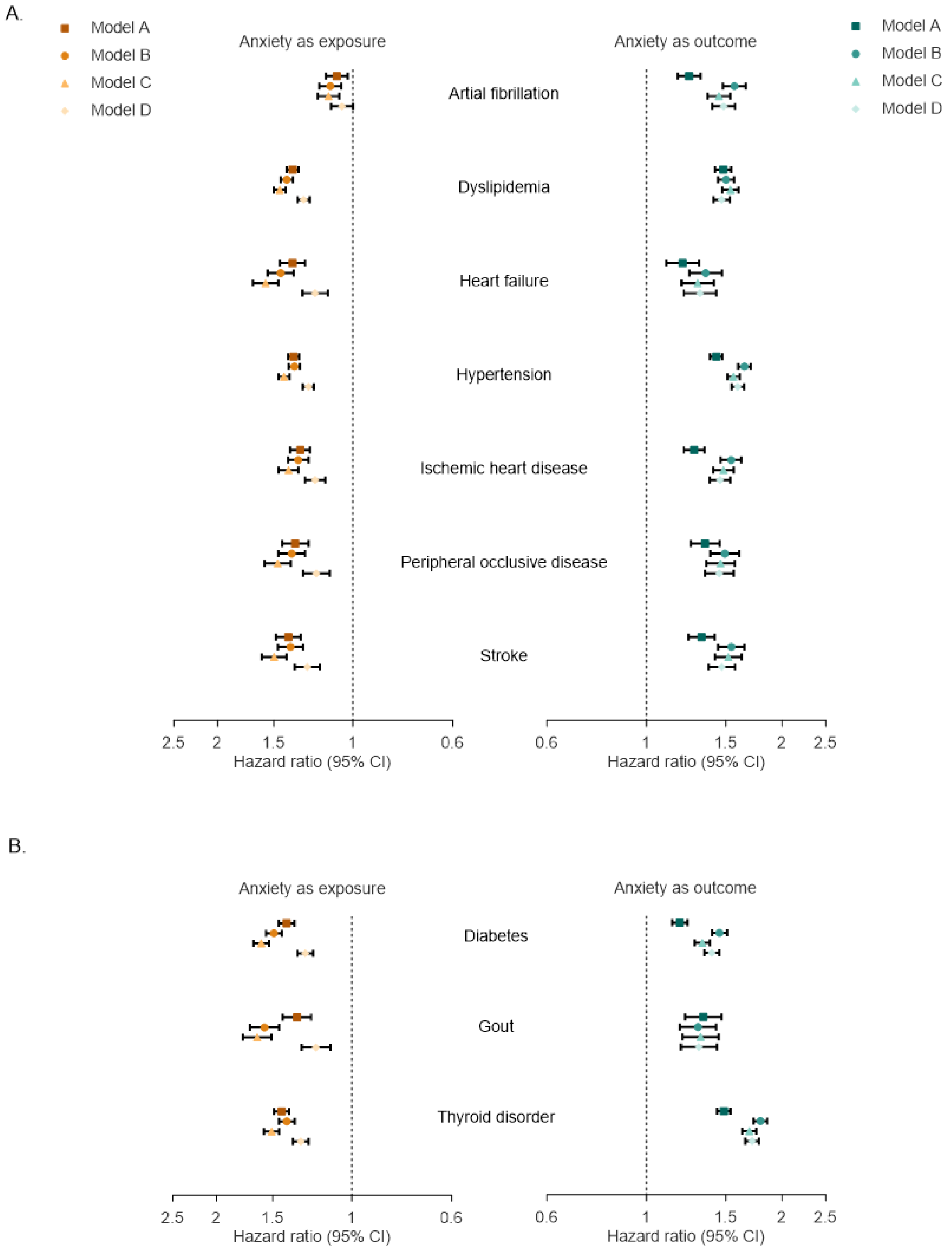

#### eFigure 5. Cox regression: A. Pulmonary, B. urogenital and C. hematological diseases

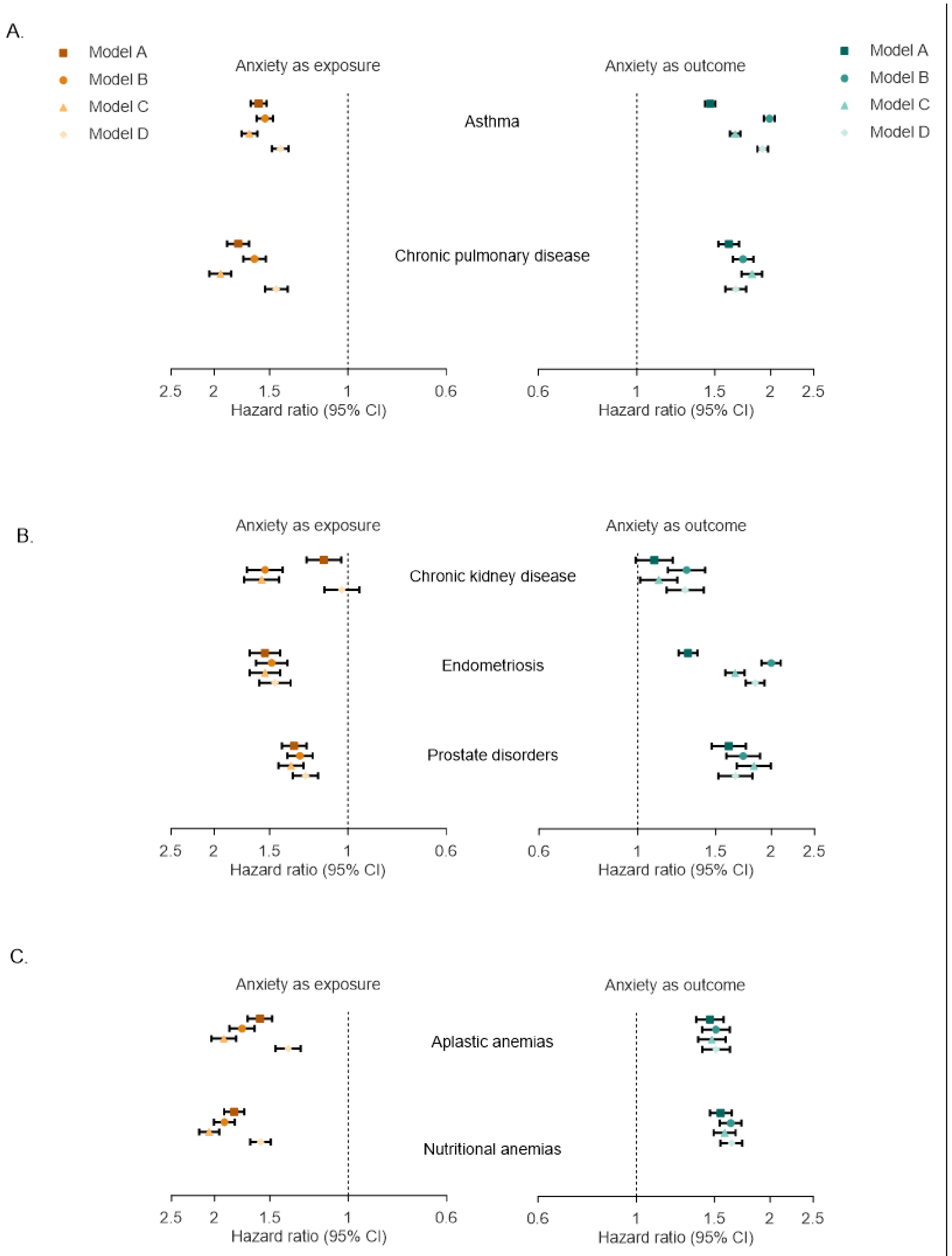

#### eFigure 6. Cox regression: A. Musculoskeletal and B. dermatological diseases

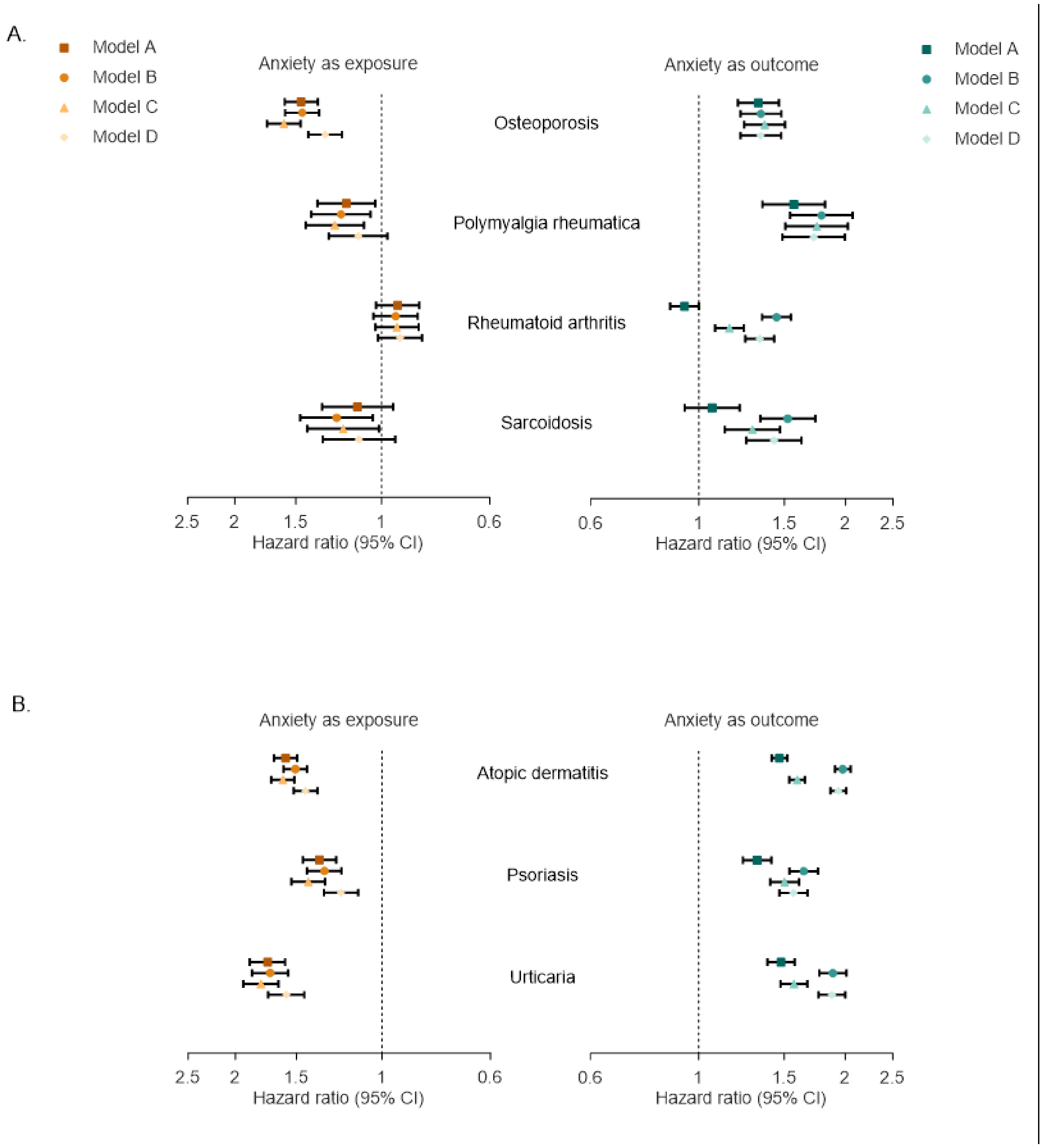

eFigures 7-9 display bidirectional hazard ratios (HR) and 95% confidence intervals (CI) obtained from time-dependent Cox proportional hazard models, using age as the underlying timescale, where (first) diagnosis of an anxiety disorder is either treated as the exposure (orange) or the outcome (teal). The HRs, shown on logarithmic scale on the y-axis, were estimated across varying lengths of exposure to the preceding condition which ranged from 0-6 months up to ≥15 years of exposure, as shown on the x-axis. The analyses were conducted across 11 different broad categorical groups of somatic diagnoses and with four different models of adjustments (Models A, B, C and D). Model A is adjusted with sex and calendar time; model C is adjusted further with somatic comorbidities diagnosed before the onset of the exposing condition and the total number of these comorbidities and model D with psychiatric comorbidities diagnosed before the onset of the exposing condition and the total number of these psychiatric comorbidities. Line of unity is shown in each panel with the dotted lines. Results from model B can be seen in Figure 3A.

#### eFigure 7. Bidirectional time-dependent Cox regression: Broad GMC categories (model A)

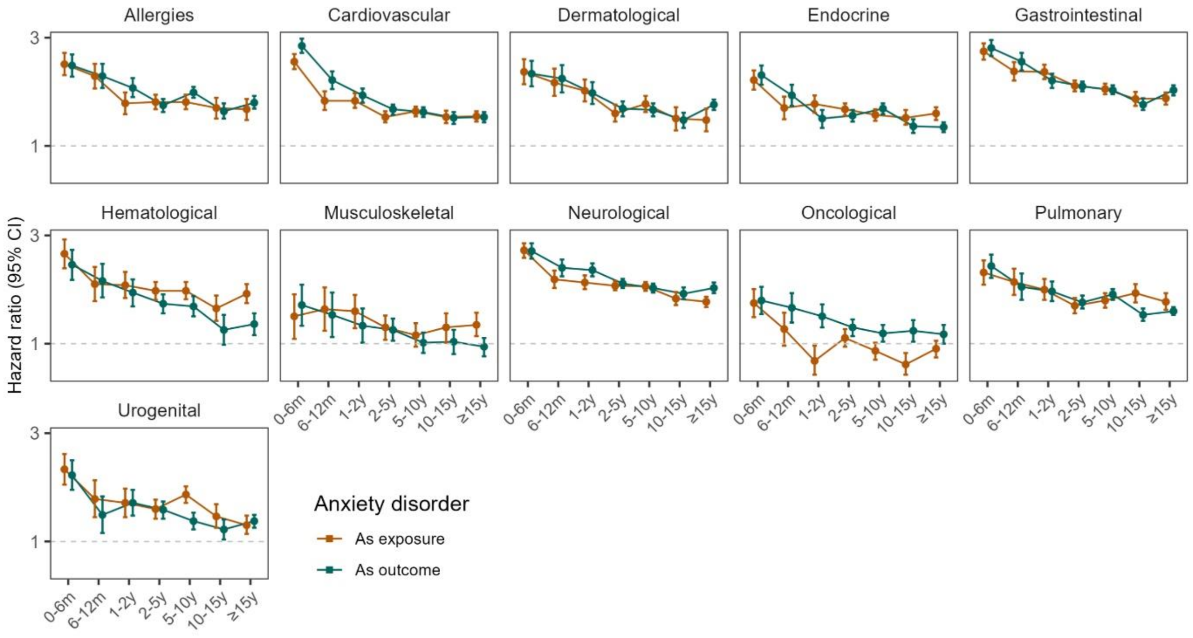

#### eFigure 8. Bidirectional time-dependent Cox regression: Broad GMC categories (model C)

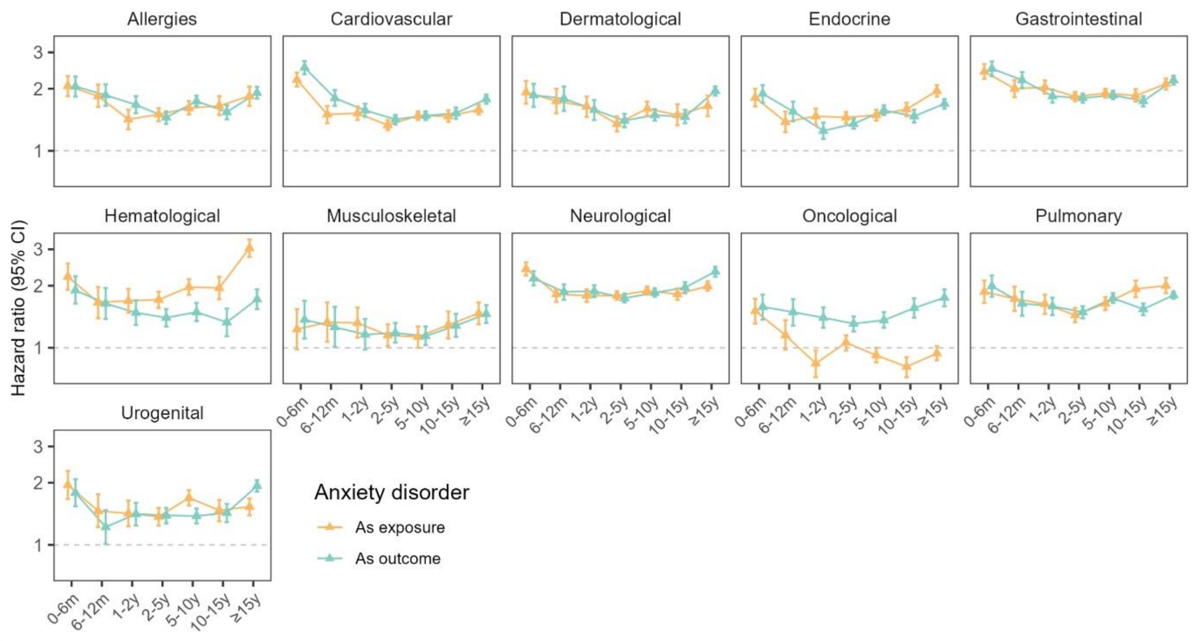


#### eFigure 9. Bidirectional time-dependent Cox regression: Broad GMC categories (model D)

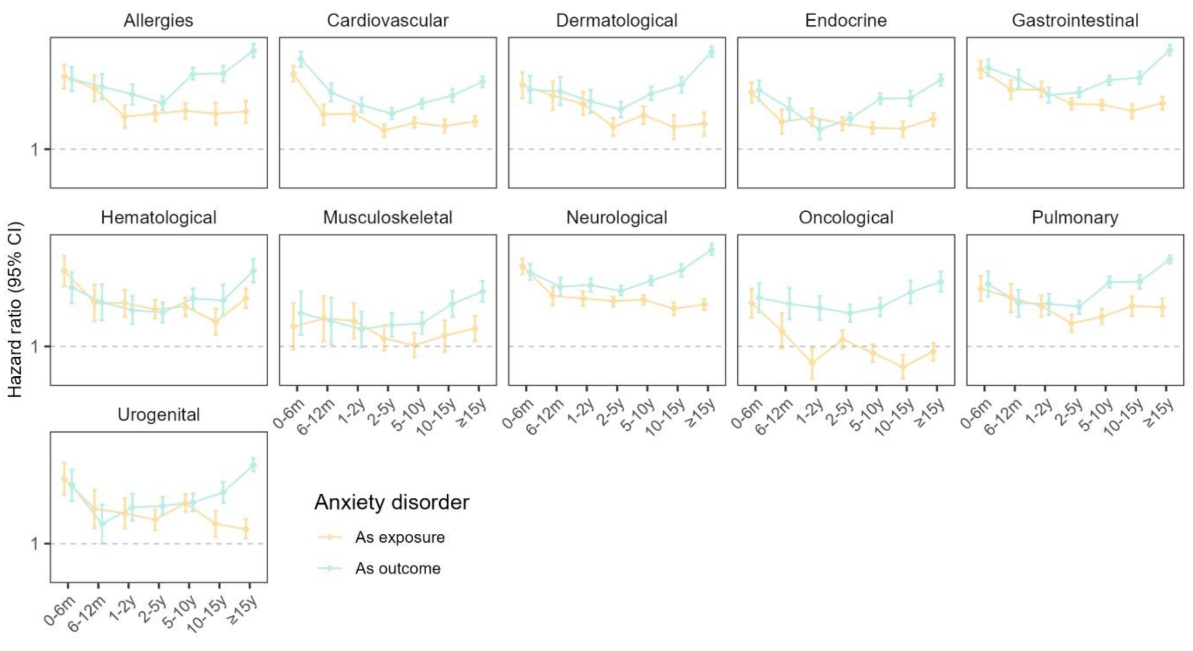

Supplementary figures 10-18 display bidirectional hazard ratios (HR) and 95% confidence intervals (CI) obtained from time-dependent Cox proportional hazard models, using age as the underlying timescale, where (first) diagnosis of an anxiety disorder is either treated as the exposure (orange) or the outcome (teal). The HRs, shown on logarithmic scale on the y-axis, were estimated across varying lengths of exposure to the preceding condition which ranged from 0-6 months up to ≥15 years of exposure, as shown on the x-axis. The analyses were conducted across 45 individual diseases and with four different models of adjustments (Models A, B, C and D). Model A is adjusted with sex and calendar time, model B is adjusted further with both somatic and psychiatric comorbid conditions diagnosed before the onset of the preceding exposing condition and the number of comorbid conditions from both of these classes, model C is adjusted further with only somatic comorbidities diagnosed before the onset of the exposing condition and the total number of somatic comorbidities and model D with only psychiatric comorbidities diagnosed before the onset of the exposing condition and the total number of psychiatric comorbidities. Line of unity is shown in each panel with the dotted lines. For gastrointestinal and neurological diseases results from the model B can be seen in Figure 3B and 3C.

**eFigure 10. Bidirectional time-dependent Cox regression: Gastrointestinal diseases (models A, C and D)**

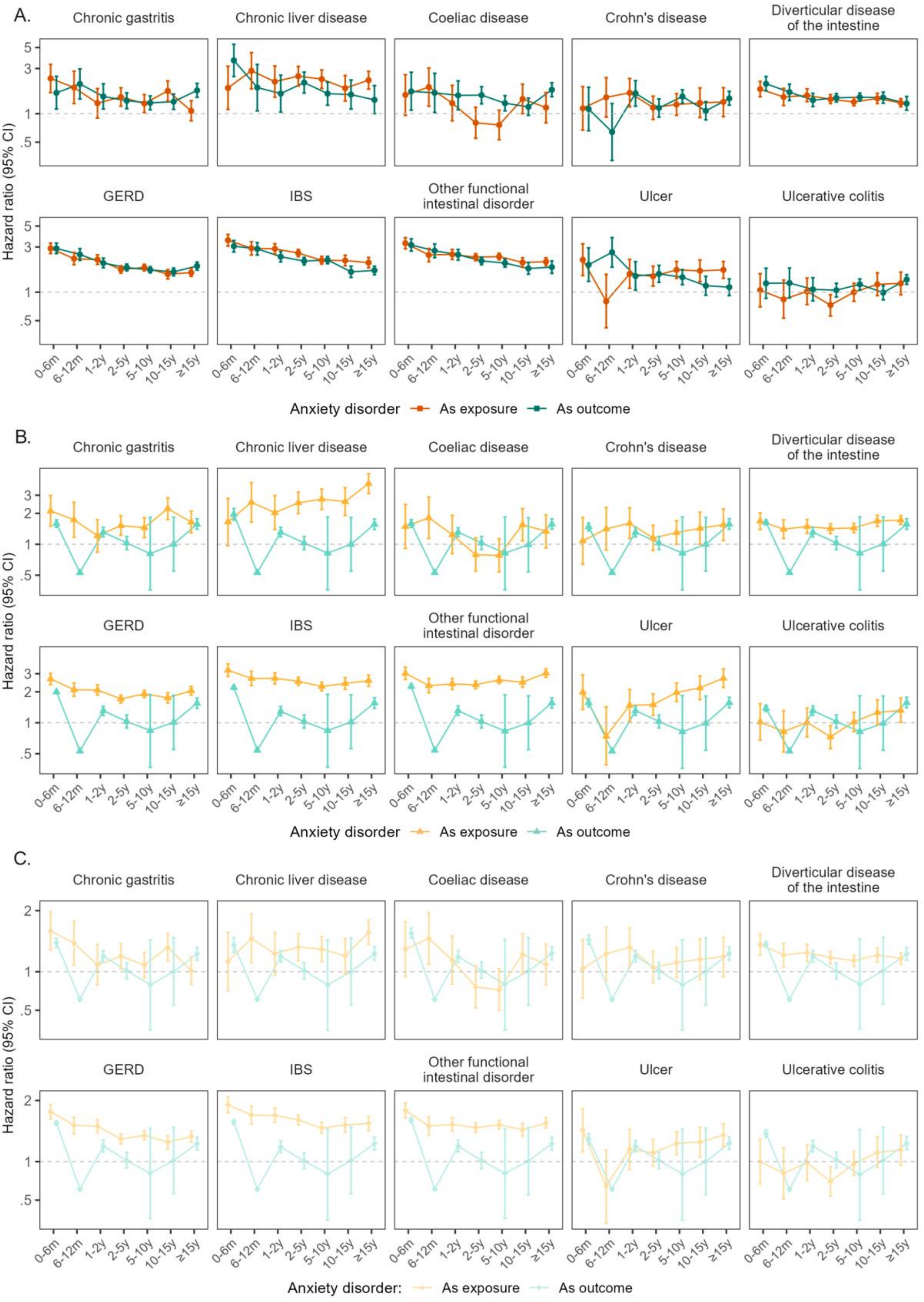

#### eFigure 11. Bidirectional time-dependent Cox regression: Neurological diseases (models A, C and D)

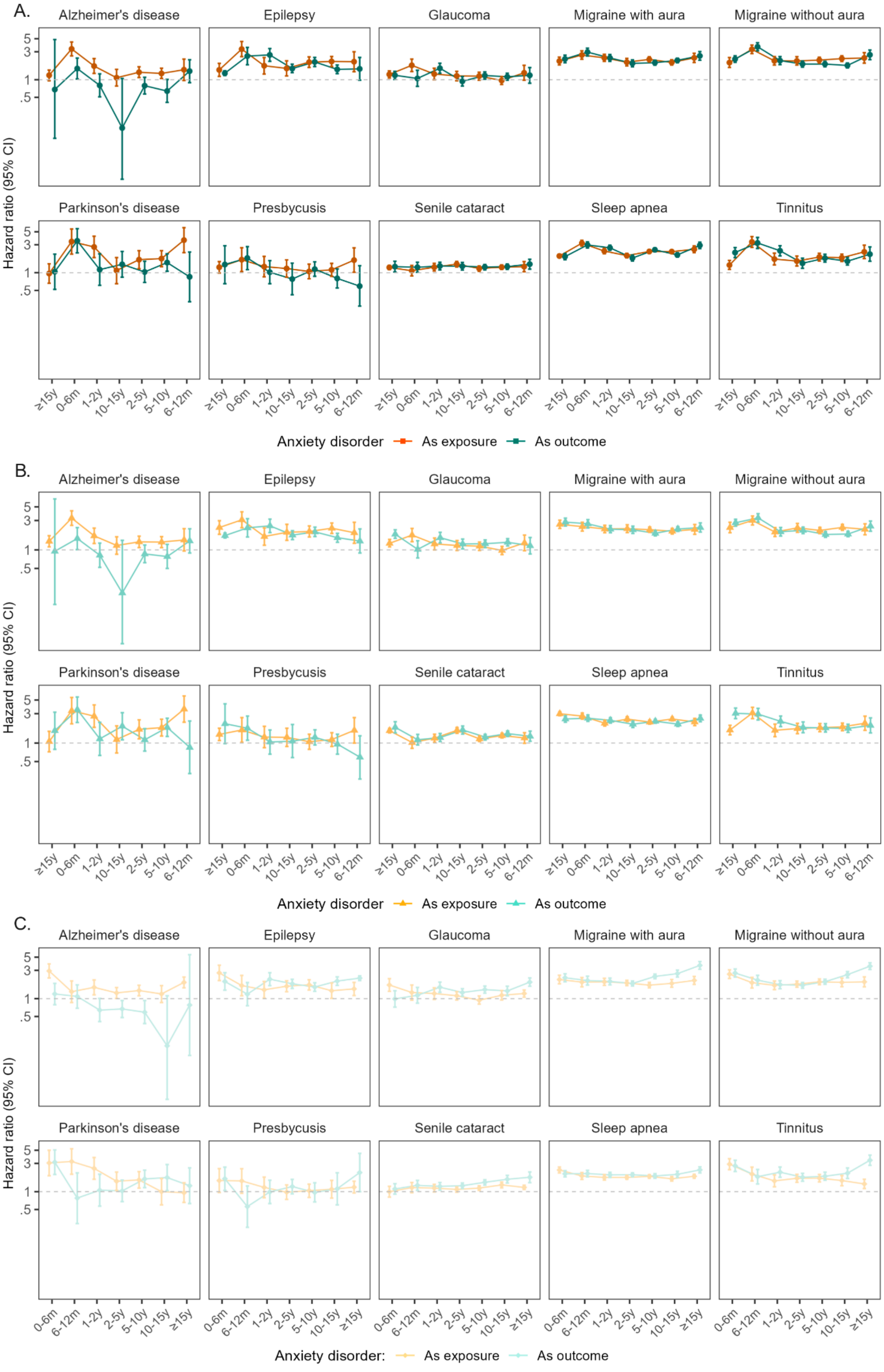

#### eFigure 12. Bidirectional time-dependent Cox regression: Cardiovascular diseases (all models)

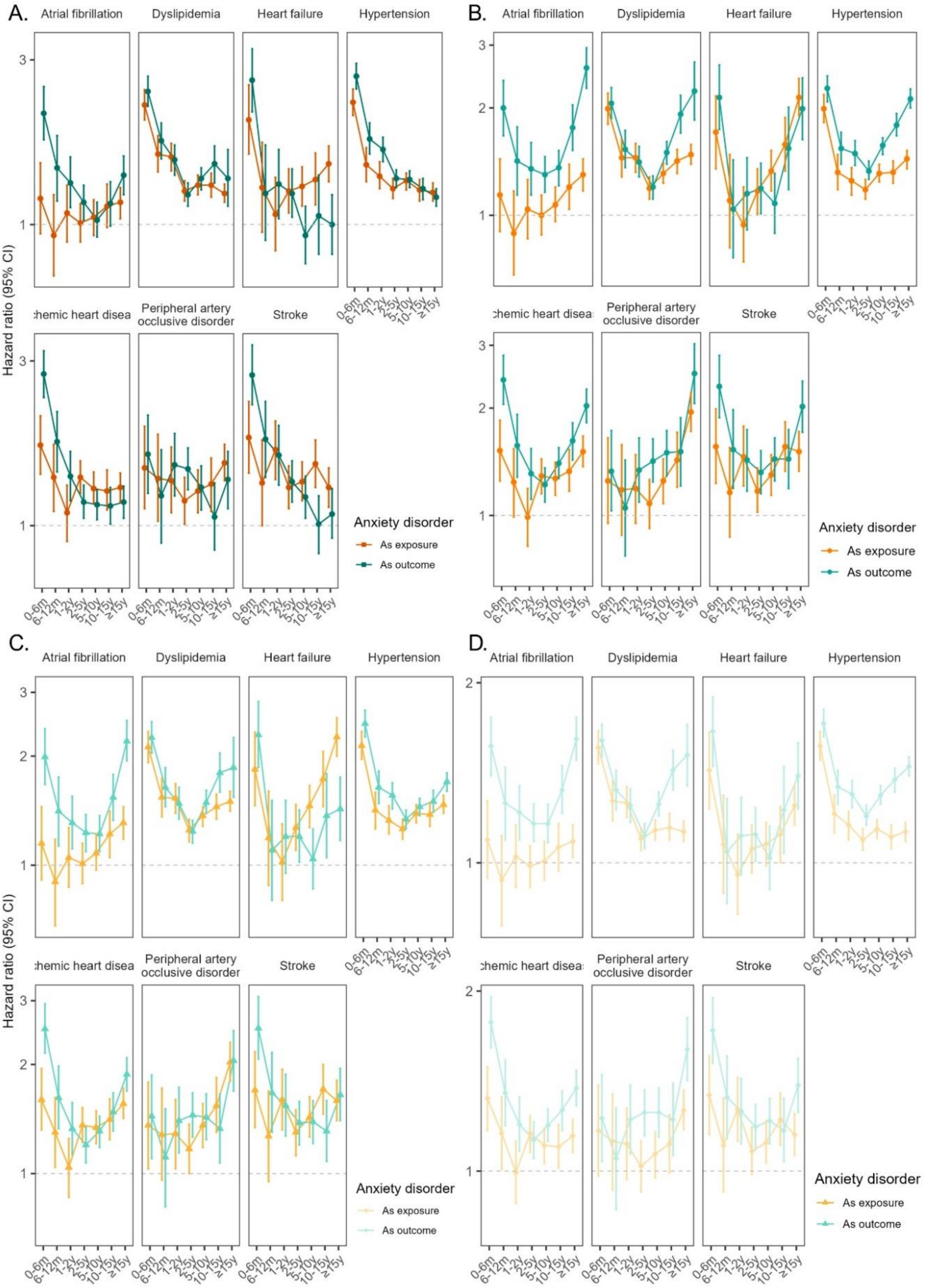

#### eFigure 13. Bidirectional time-dependent Cox regression: Endocrine diseases (all models)

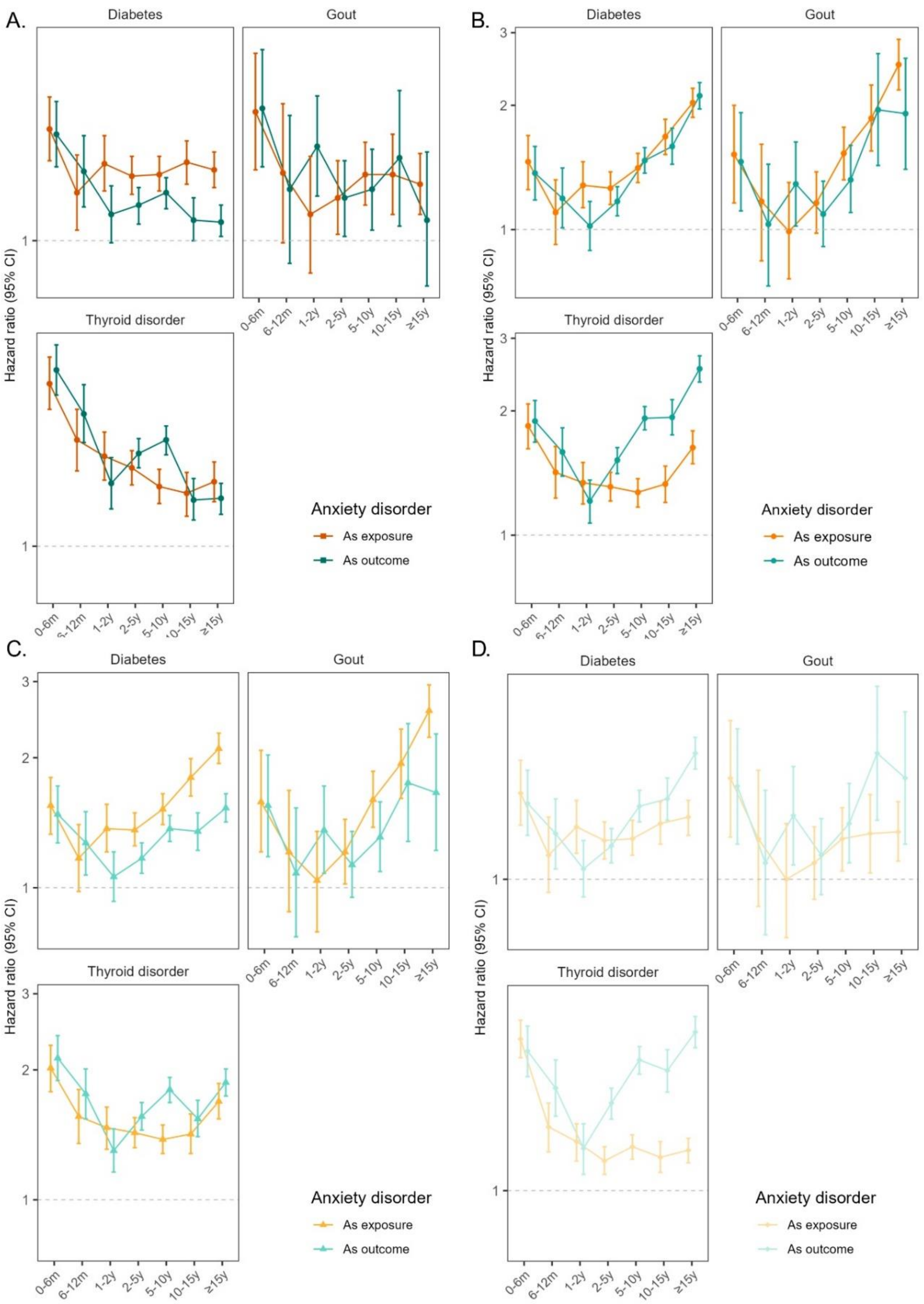

#### eFigure 14. Bidirectional time-dependent Cox regression: Pulmonary diseases (all models)

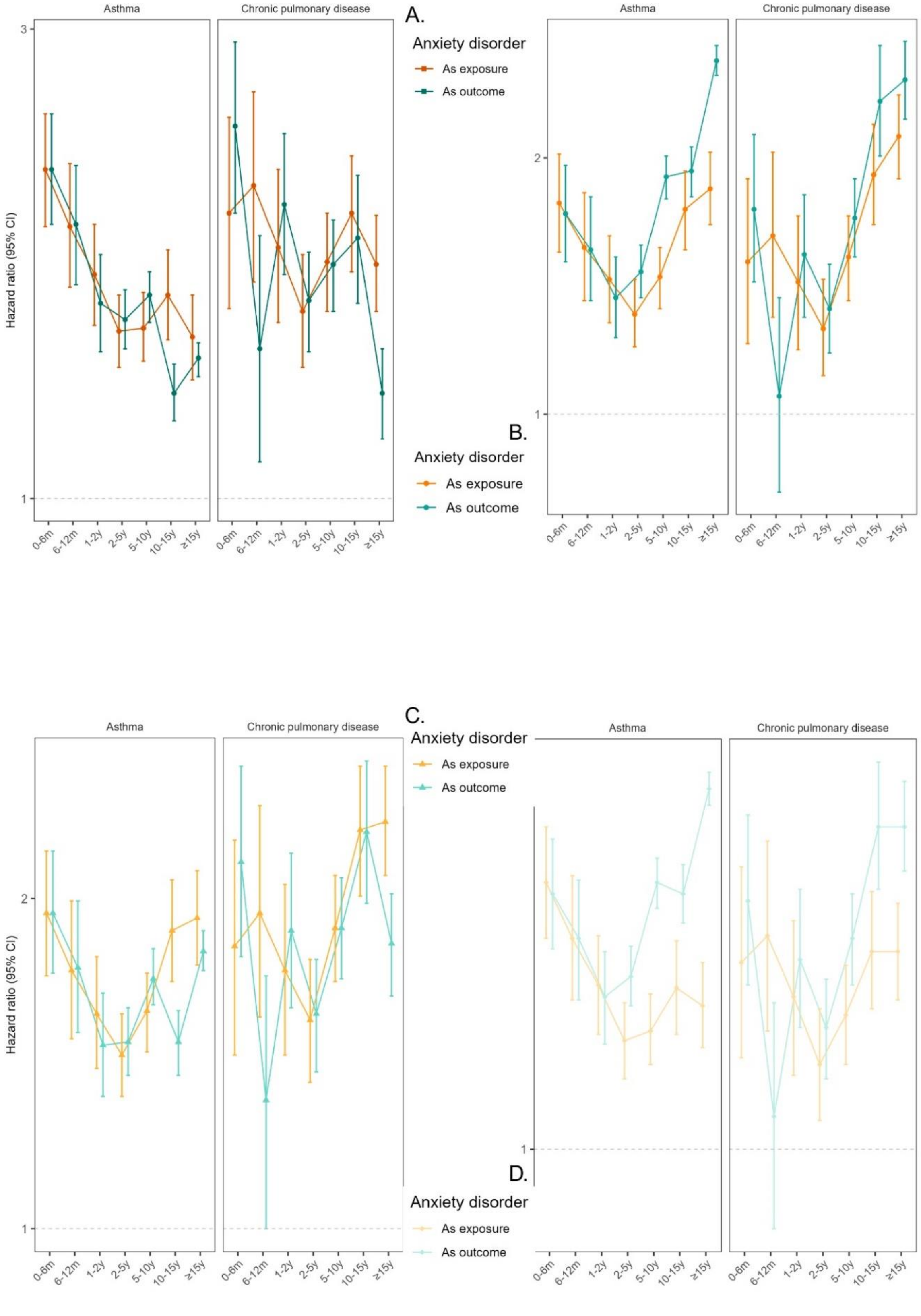

#### eFigure 15. Bidirectional time-dependent Cox regression: Urogenital diseases (all models)

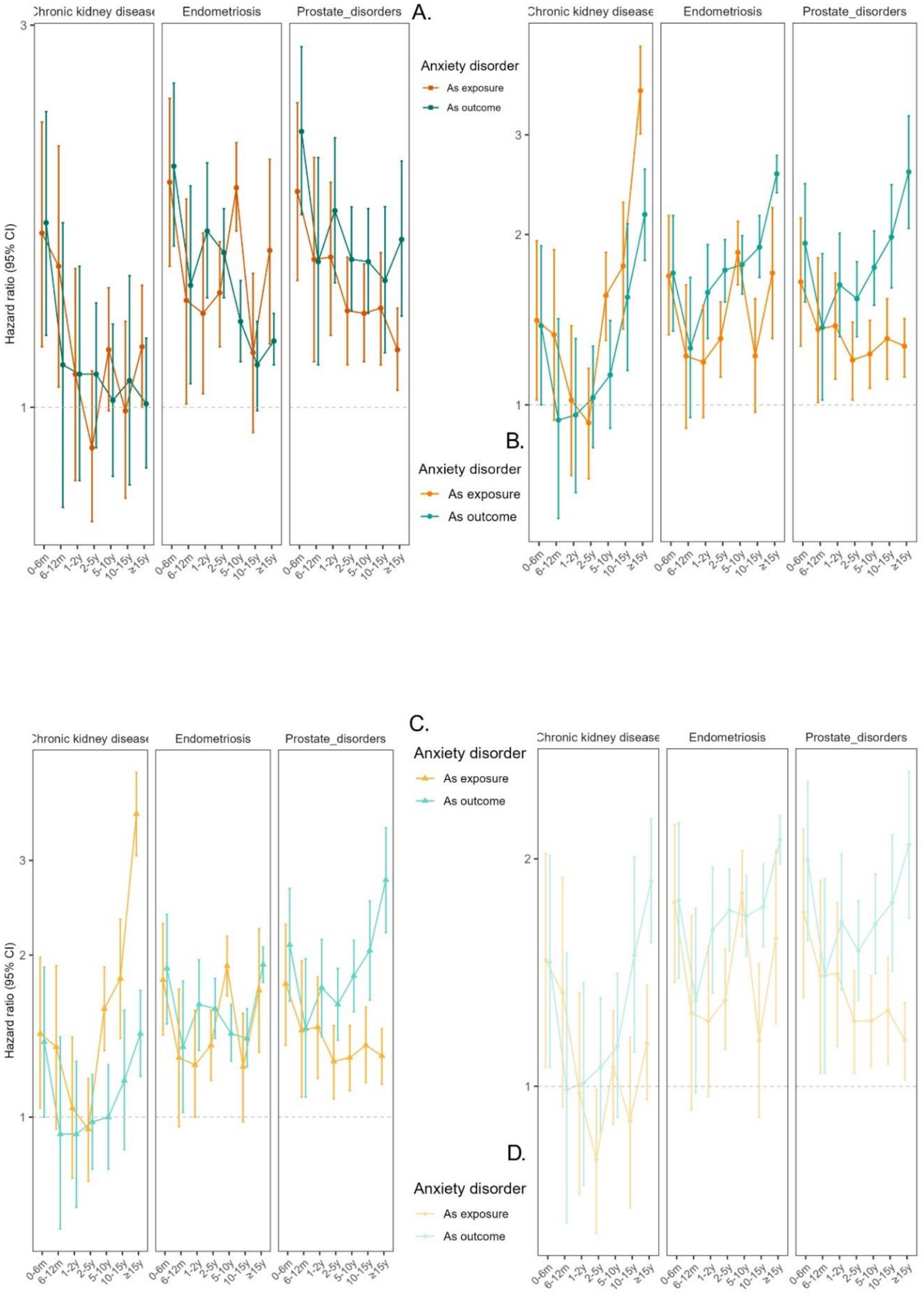

#### eFigure 16. Bidirectional time-dependent Cox regression: Musculoskeletal diseases (all models)

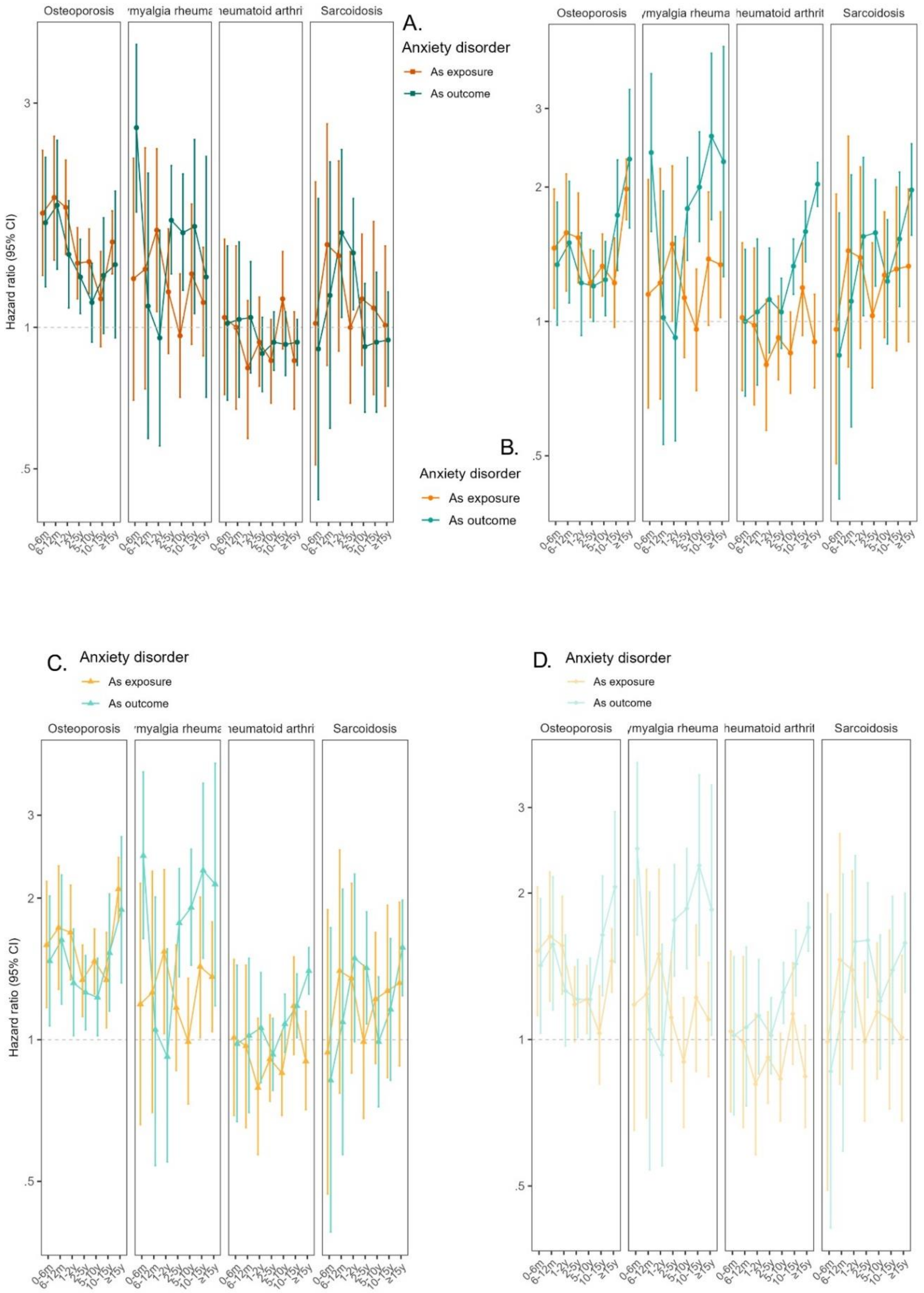

#### eFigure 17. Bidirectional time-dependent Cox regression: Hematological diseases (all models)

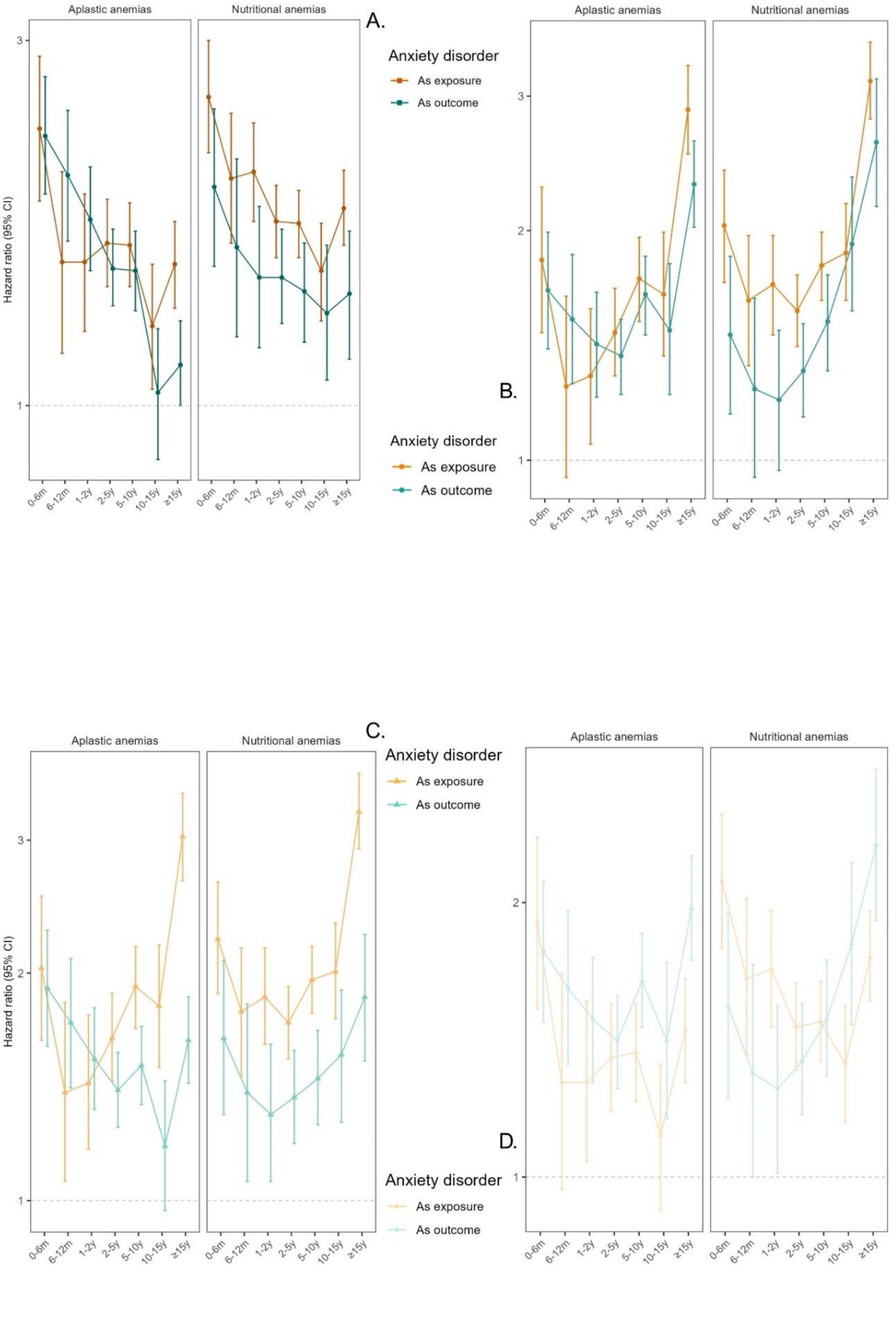

#### eFigure 18. Bidirectional time-dependent Cox regression: Dermatological diseases (all models)

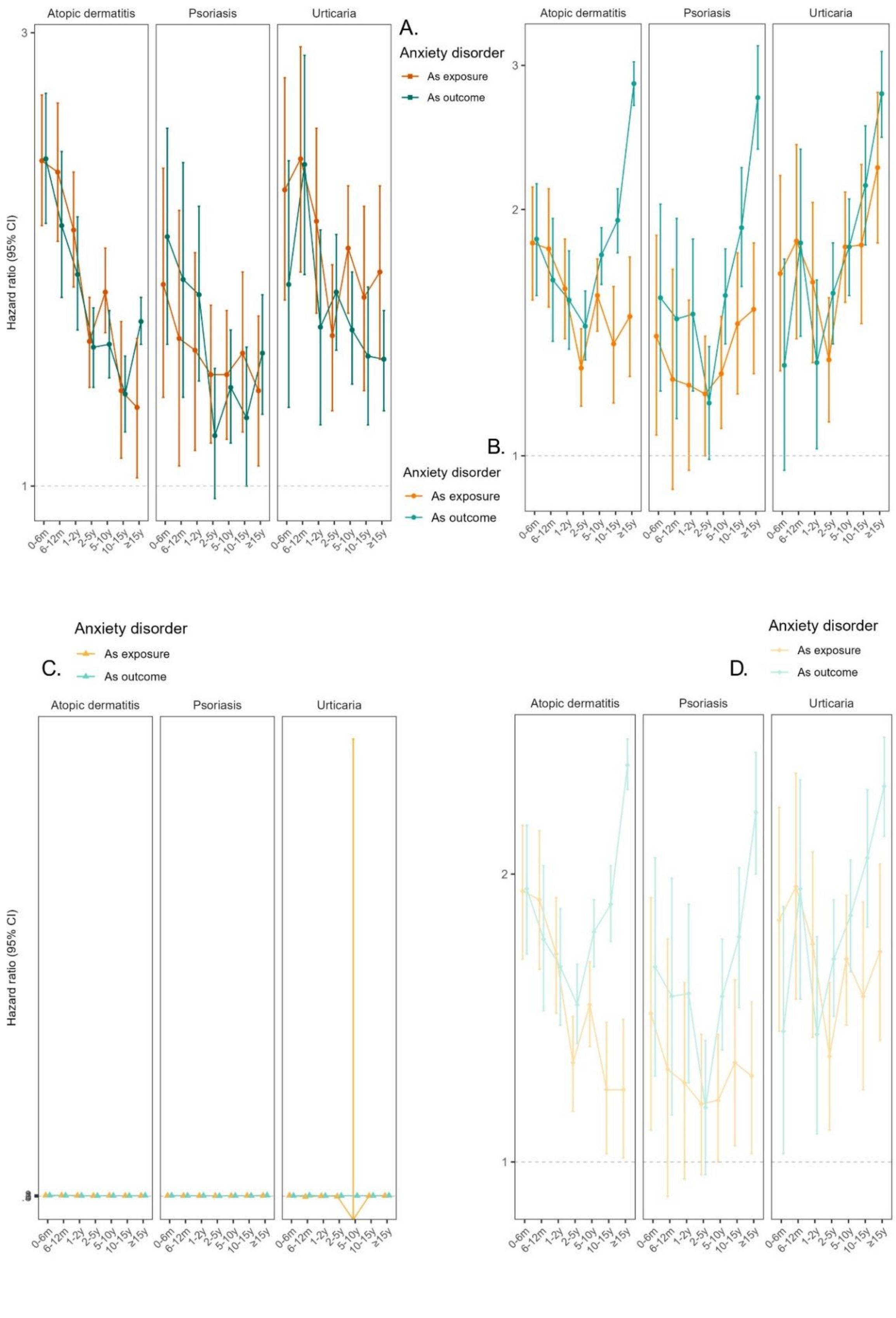

#### eFigure 19. Initial genetic correlation results

Genetic correlation ($r_g$)[20] between different anxiety disorder definitions (x-axis) and FinnGen custom psychiatric disorders and general medical conditions including cardiovascular, endocrinological, pulmonary, allergic, dermatological, hematological, oncological and urogenital diseases (eTables 1,2) (y-axis). We used six different anxiety disorder definitions: custom FinnGen GWAS summary-statistics with curated five anxiety disorder phenotypes from hospital and primary care data (Anxiety wide, Anxiety narrow, Anxiety strict, Anxiety with major depressive disorder (MDD) excluded from controls, and Anxiety with MDD excluded from both cases and controls (all), see eTable 3 for detailed phenotype definitions) and the PGC (Psychiatric Genomics Consortium) anxiety disorder meta-analysis leave-one out FinnGen summary statistics[3]. Created with BioRender.com.

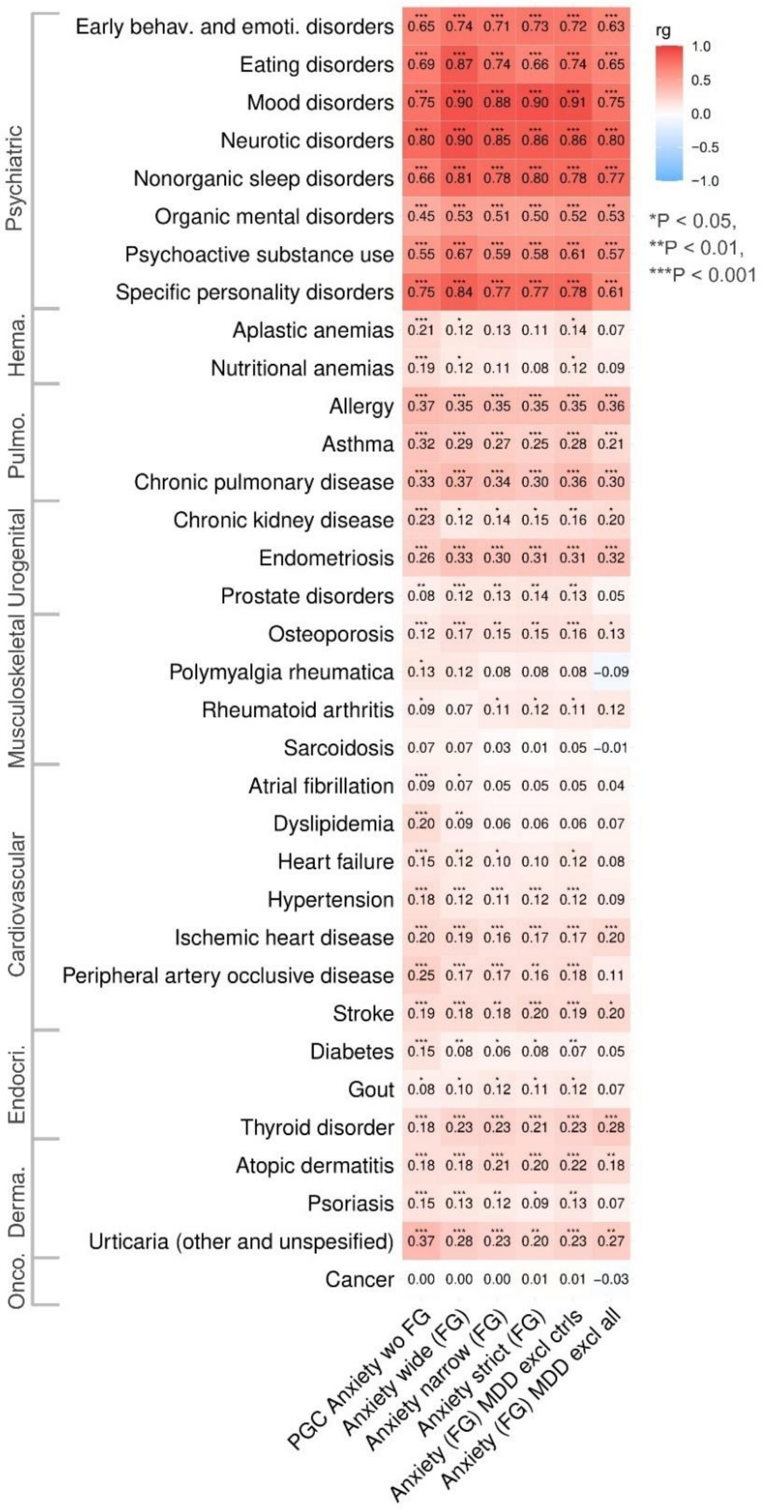

#### eFigure 20. Genetic correlation with publicly available data and FinnGen endpoints

Genetic correlation ($r_g$)[20] between different anxiety disorder definitions (x-axis) and publicly available GWAS summary statistics (eTable 5)[21–26,28–30,33,43] and FinnGen endpoints (eTable 6) of general medical conditions (y-axis). We used six different anxiety disorder definitions: custom FinnGen GWAS summary-statistics with curated five anxiety disorder phenotypes from hospital and primary care data (Anxiety wide, Anxiety narrow, Anxiety strict, Anxiety with major depressive disorder (MDD) excluded from controls, and Anxiety with MDD excluded from both cases and controls (all), see eTable 3 for detailed phenotype definitions) and the PGC (Psychiatric Genomics Consortium) anxiety disorder meta-analysis leave-one out FinnGen summary statistics[3]. Created with BioRender.com.

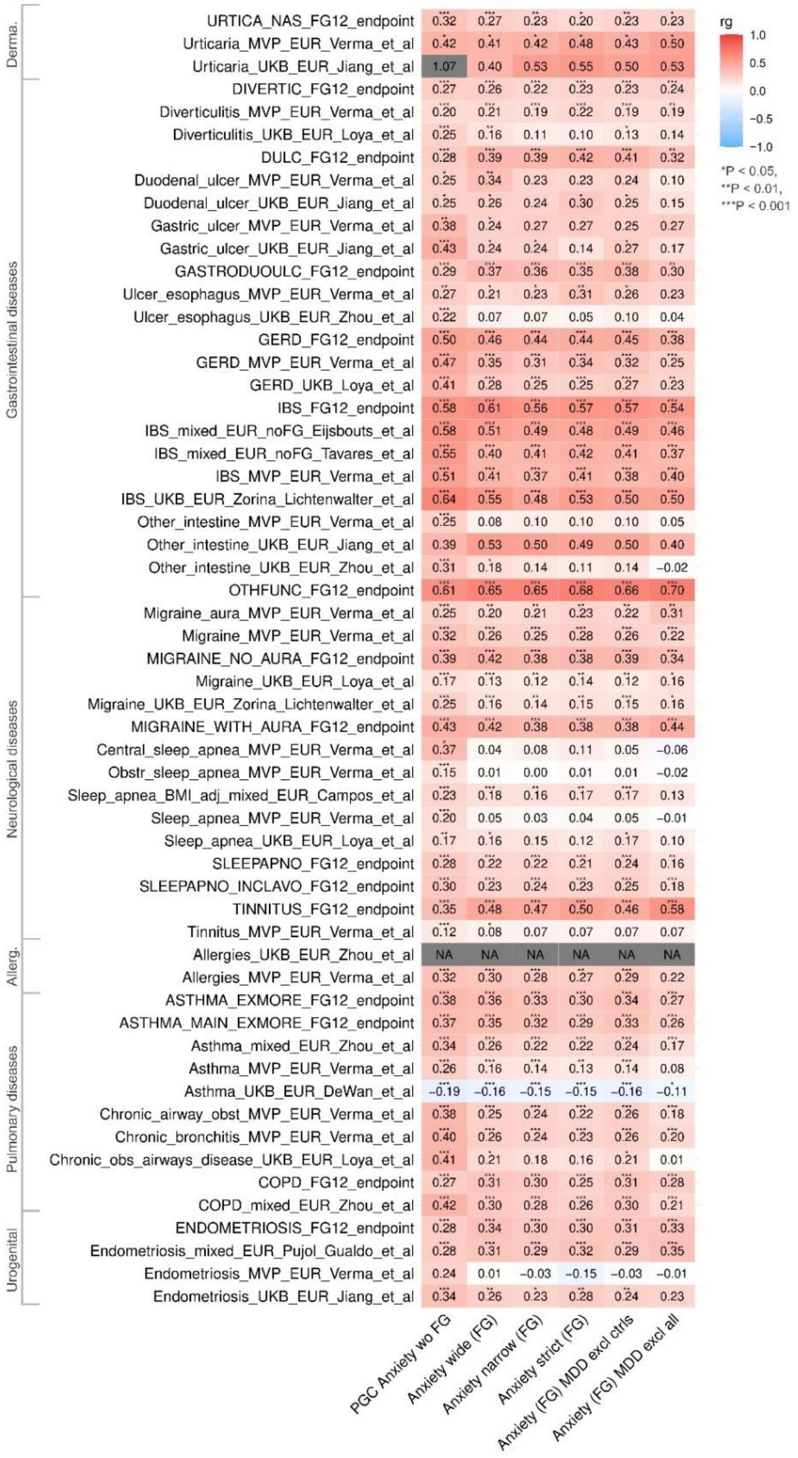

#### eFigure 21. CPASSOC PGC Anxiety and meta-analysis of IBS

Manhattan plots (-$\log_{10}$ P-value on y-axis and chromosomes on the x-axis) of CPASSOC pleiotropy analysis[37] between the PGC (Psychiatric Genomics Consortium) anxiety disorder meta-analysis leave-one out FinnGen[3] and meta-analysis of irritable bowel syndrome (IBS), **A.** SHom (Homogenous effective model) and **B.** SHet (Heterogenous effective model). Top 5 lead variants by P-value are marked with their nearest genes (by genomic position). Created with BioRender.com.

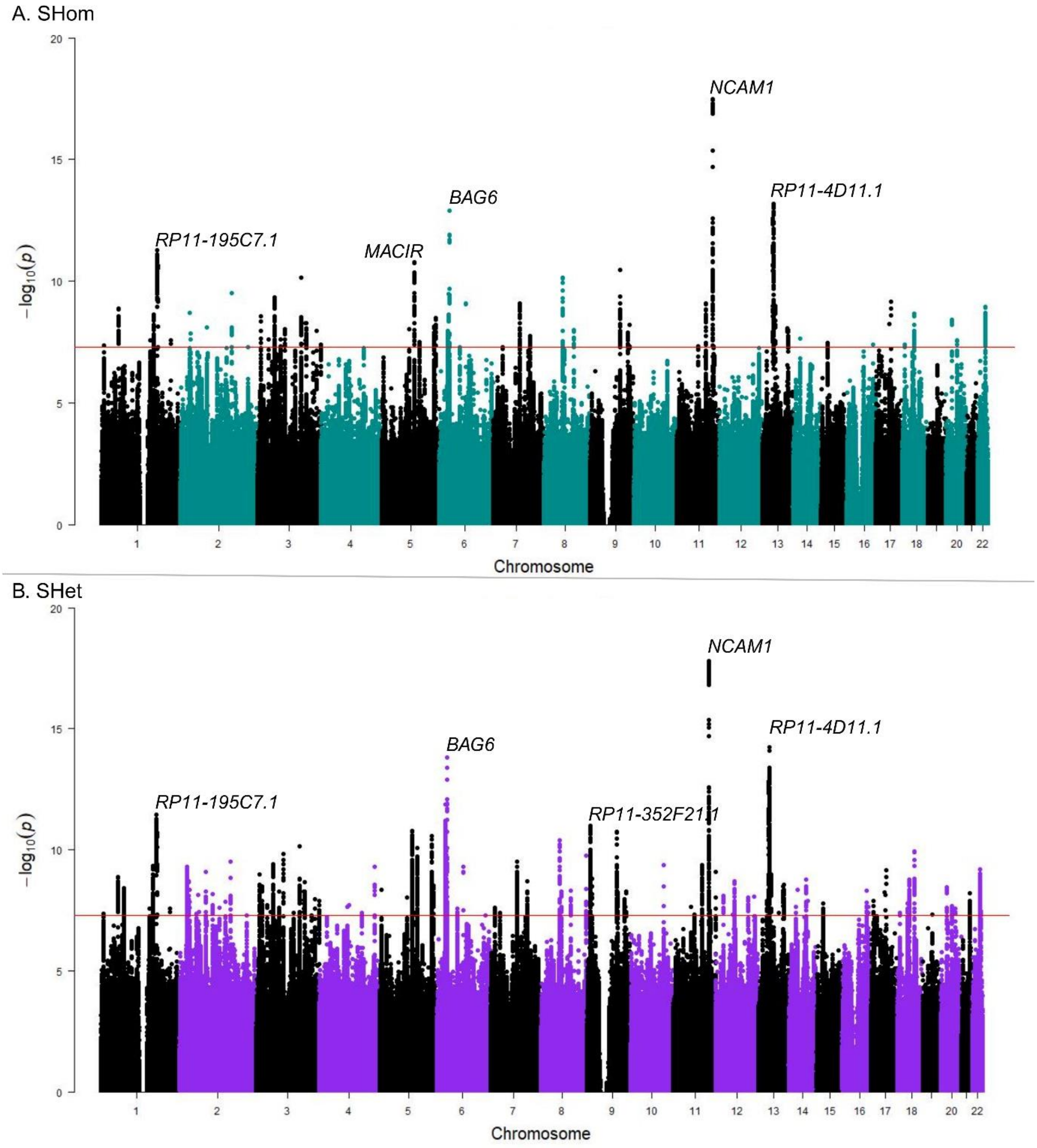

#### eFigure 22. CPASSOC PGC Anxiety and meta-analysis of GERD

Manhattan plots (-$\log_{10}$ P-value on y-axis and chromosomes on the x-axis) of CPASSOC pleiotropy analysis[37] between the PGC (Psychiatric Genomics Consortium) anxiety disorder meta-analysis leave-one out FinnGen[3] and meta-analysis of gastro-esophageal disease (GERD), **A.** SHom (Homogenous effective model) and **B.** SHet (Heterogenous effective model). Top 5 lead variants by P-value are marked with their nearest genes (by genomic position).

A. SHom

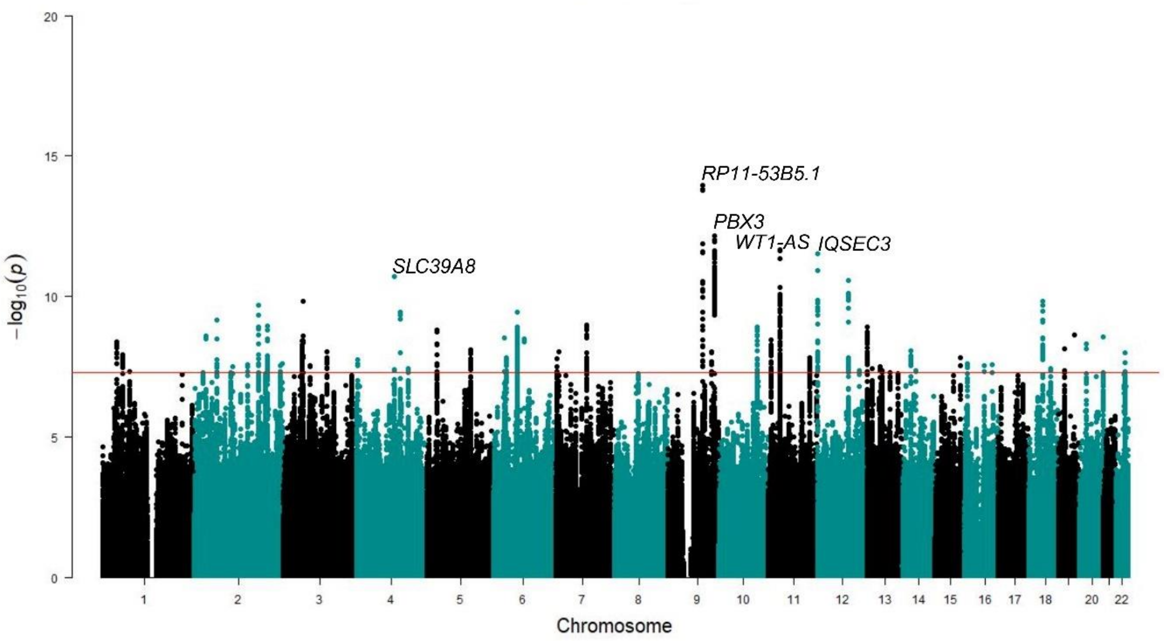


B. SHet

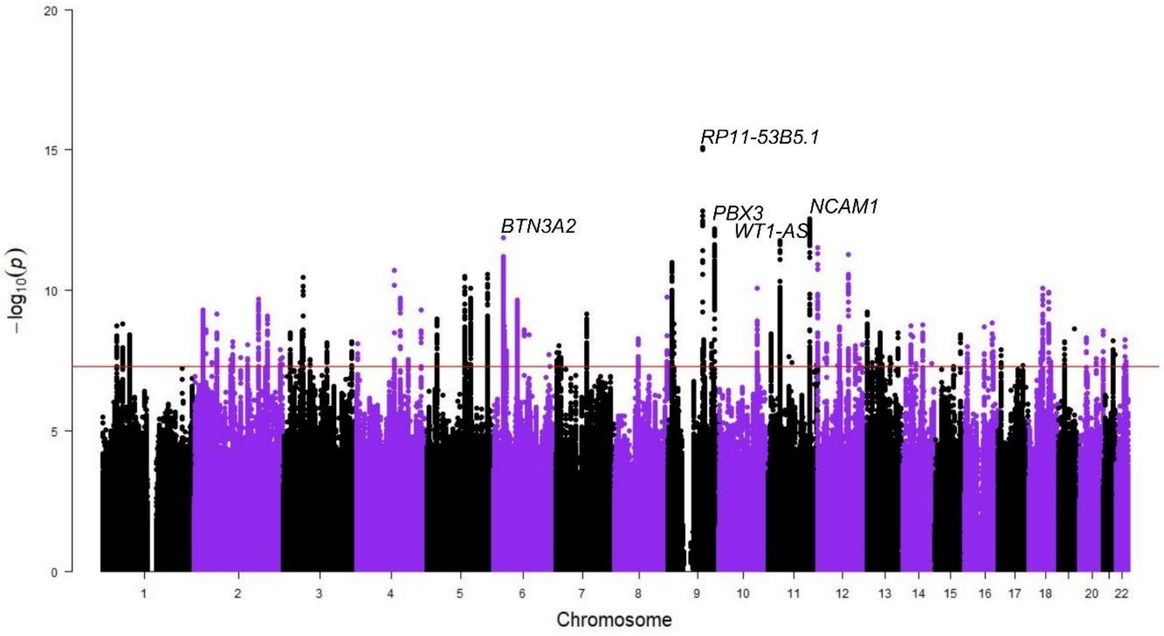

#### eFigure 23. Tissue-level gene expression

Average of gene expression value per label (e.g. tissue types or developmental stage, the GTEx project[42]) of the shared genes between **A.** anxiety disorders and IBS, and **B.** anxiety disorders and GERD (eTable 17). Cells with red represent higher expression across labels and genes. IBS: Irritable bowel syndrome, GERD: gastro-esophageal reflux.

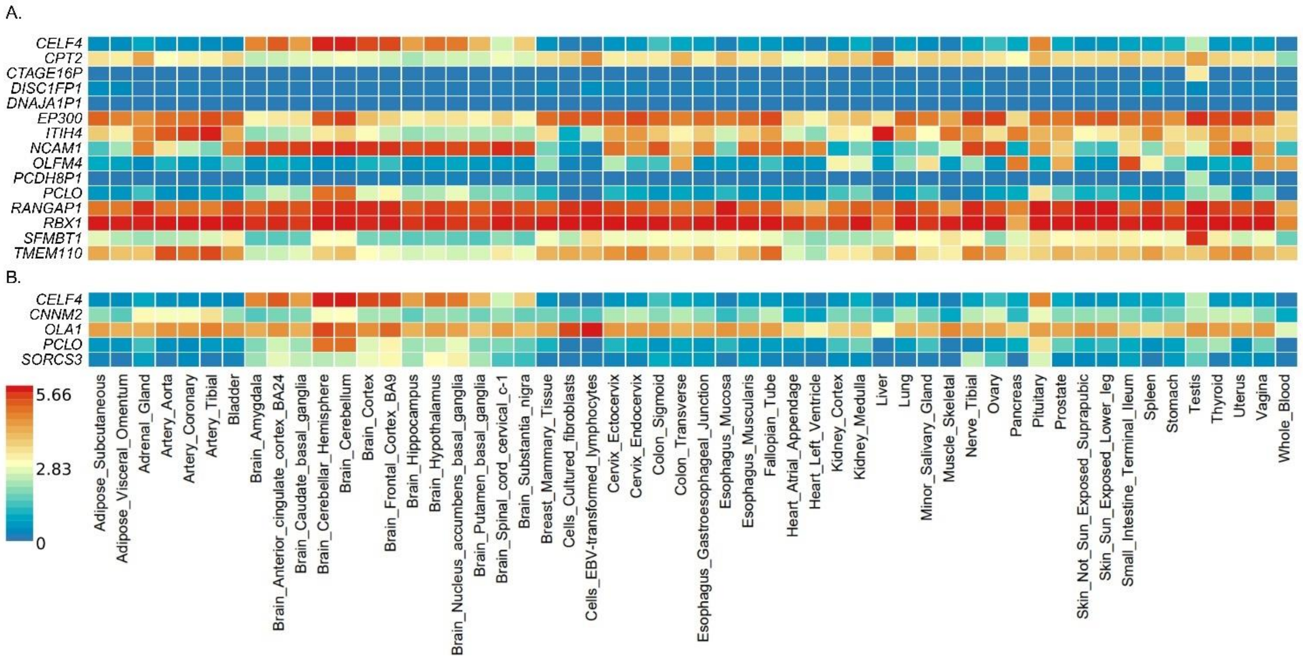
